# Vulnerabilities in child wellbeing among primary school children: a cross-sectional study in Bradford, UK

**DOI:** 10.1101/2021.01.10.21249538

**Authors:** Kate E Pickett, Mildred Ajebon, Bo Hou, Brian Kelly, Philippa K Bird, Josie Dickerson, Katy Shire, Mark Mon-Williams, Neil Small, Rosemary McEachan, John Wright, Deborah A Lawlor

## Abstract

**Objective:** To describe the prevalence of factors related to wellbeing among primary school children in a deprived multi-ethnic community.

**Design and participants:** Cross-sectional survey of 15,641 children aged 7-10 years in Born in Bradford’s Primary School Years study: whole-classroom samples in 89 Bradford primary schools between 2016 and 2019.

**Main outcome measures:** Prevalence estimates by ethnicity (%, 95% CI) of single and multiple vulnerabilities in child wellbeing within and across four domains (home, family, relationships; material resources; friends and school; subjective wellbeing).

**Results:** Only 10% of children have no vulnerabilities in any domain of wellbeing; 10% have one or more vulnerabilities in all four domains. The highest prevalence estimates were for being bullied some or all of the time (52.7%, 51.9 to 53.4%), keeping worries to oneself (31.2%, 30.5 to 31.9%), having no park near home (30.8%, 30.1 to 31.5%) and worrying all the time about how much money their family has (26.3%, 25.6 to 27%).

Boys were consistently significantly more likely than girls to report all of the vulnerabilities in the Home, Family and Family Relationships domain, and the majority of indicators in the other domains, and in all domains except Friends and School, boys were significantly more likely to have at least one vulnerability. Girls were significantly more likely to report not having many friends (16.7%, 95% CI: 15.9 to 17.6% vs. 12.5%, 95% CI: 11.8 to 13.2%), being bullied some or all of the time (55.8%, 95% CI: 54.7 to 56.9% vs. 49.7%, 95% CI: 48.6 to 50.8%) and feeling left out all the time (12.1%, 95% CI: 11.4 to 12.8%) vs. 10.3%, 95% CI: 9.7 to 11.0%).

Variations in vulnerabilities by ethnicity were complex, with children in Black, Asian and Minority Ethnic groups sometimes reporting more vulnerabilities and sometimes fewer than White British children. For example, compared to children of Pakistani heritage, White British children were more likely to say that their family never gets along well (6.3%,5.6 to 7.1% vs. 4.1%,3.6 to 4.6%) and to have no access to the internet at home (22.3%,21 to 23.6% vs. 18%,17 to 18.9%). Children with Pakistani heritage were more likely than White British children to say they had no park near their home where they can play with friends (32.7%,31.6 to 33.9% vs. 29.9%,28.6 to 31.3%), to report not having three meals a day (17.9%,16.9 to 18.8% vs. 11.9%,10.9 to 12.9%) and to worry all the time about how much money their families have (29.3%,28.2 to 30.3%) vs. 21.6%,20.4 to 22.9%). Gypsy/Irish Traveller children were less likely than White British children to say they were bullied some or all of the time (42.2%,35.4 to 49.4% vs. 53.8%,52.3 to 55.3%), but more likely to say they were mean to others all the time (9.9%,6.3 to 15.2% vs. 4%,3.5 to 4.7%) and can never work out what to do when things are hard (15.2%,10.6 to 21.2% vs. 9%, 8.2 to 9.9%).

We considered six vulnerabilities to be of particular concern during the current Covid-19 pandemic and associated national and local lockdowns: family never gets along well together; no garden where child can play; no nearby park where they can play; not having 3 meals a day; no internet at home; worried about money all the time. Pre-pandemic, 37.4% (36.6 to 38.3%) of Bradford children had one of these vulnerabilities and a further 29.6% (28.9 to 30.4%) had more than one.

**Conclusions:** Although most primary school children aged 7-10 in our study have good levels of wellbeing on most indicators across multiple domains, fewer than 10% have no vulnerabilities at all, a worrying 10% have at least one vulnerability in all the four domains we studied and two thirds have vulnerabilities of concern during the Covid-19 lockdowns.

## Introduction

In 2007, the UK ranked bottom of 21 countries in UNICEF’s first report on child wellbeing in rich countries,^1^ and from 2009 through 2019 the Good Childhood inquiry^2^ and subsequent reports from the Children’s Society^3^ have found increasing levels of unhappiness and mental distress among children and young people in the UK. The State of the Nation 2019 report on children and young people’s wellbeing suggested a need to “understand wellbeing across different groups” and to “use a range of measures to understand their experience.”^4^

Wellbeing declines as children and young people grow older^4^ but little is known about wellbeing in primary school aged children in the UK as previous studies, including the Good Childhood surveys, Understanding Society and HeadStart, include children over 10 years old only. The impact of ethnicity on child wellbeing at any age is unclear and most previous UK studies are unable to examine wellbeing across a wide range of ethnicities.^4^ To provide evidence to meet these knowledge gaps, we examined child wellbeing among primary school children, aged 7 to 10 years, in a city in the North of England. Bradford is the fifth largest metropolitan district in England with one of the youngest and most ethnically diverse populations.^5^ Bradford also has some of the highest levels of poverty and ill-health in England; almost a quarter of Bradford children live in poverty and 24% are obese at age 10/11.^6^

The global Covid-19 pandemic and restrictions, such as lockdowns that closed schools, heightened concerns about child wellbeing across the world. The prevalence of pre-existing child vulnerabilities and risk factors that could potentially require mitigation and targeted intervention during and after the pandemic has been a concern, particularly in poor, ethnically diverse urban settings.^7^ The aim of this paper is to describe the prevalence of vulnerabilities in child wellbeing by ethnicity and gender in a deprived community. Whilst this research is undertaken in the city of Bradford and has been used to support local policy during the COVID-19 pandemic,^8^ we believe the results have wider relevance for deprived multi-ethnic urban settings.

## Methods (for STROBE statement see Supplementary Materials: 1)

### Setting and Participants

Born in Bradford (BiB) is a cohort study of 13,500 children born between 2007-2011.^9^ Between 2016 and 2019, BiB administered the Primary School Years study as a cross-sectional survey of wellbeing of children aged 7-10 years in 89 Bradford schools (Figure 1). We surveyed whole classrooms, including children previously recruited to the ongoing BiB cohort study as well as non-BiB children. The study protocol and detailed methods of school recruitment are described elsewhere.^10 11^ Schools were given information sheets and opt-out consent forms to give to parents of children in eligible year groups ahead of the school visit; one school asked for opt-in consent to be used and this was accommodated. Fewer than 5% of parents opted out of the study.

**Figure 1:**
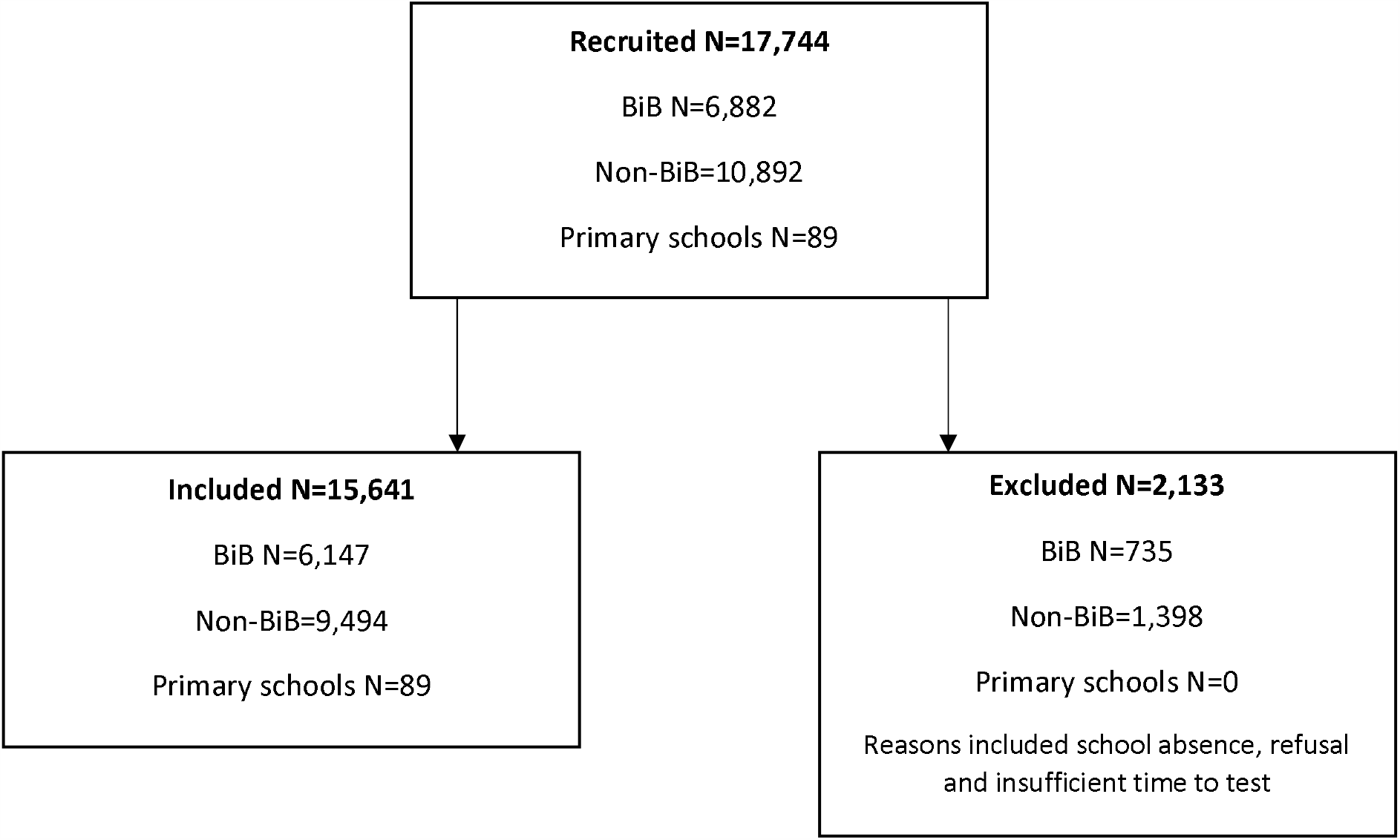
Recruitment of schools, Born in Bradford cohort members and non-Born in Bradford children to the BiB Primary School Years wellbeing survey.

### Design and Procedures

Ethical approval was obtained from the NHS Health Research Authority’s Yorkshire and the Humber - Bradford Leeds Research Ethics Committee (reference: 16/YH/0062) on the 24th March 2016.

The wellbeing survey (“Me and My Life”) was developed for this study (see Supplementary Materials: 2), drawing on questions asked of children in the Millennium Cohort Study,^12^ the International Survey of Children’s Wellbeing^13^ and the Avon Longitudinal Study of Parents and Children.^14^ The survey aimed to assess children’s wellbeing in multiple domains, including: happiness, health, material wellbeing, relationships with family and friends, school experience, neighbourhood, aspirations and acculturation. ‘Think aloud’ testing with a small group of children was used to check understanding and face validity. During administration, whole classrooms completed the survey at the same time, supported by three research facilitators.

Schools provided class lists, including date of birth, gender and ethnicity. Child ethnicity is reported by parents on registration with a school. The ethnicity information provided from school records contained 192 categories. These were re-coded into 18 broad groups in line with the 2011 census categorisation of ethnic groups in the UK and for the analyses presented here collapsed into 10 categories: Pakistani, Bangladeshi, Indian, Black/Black British, White British, Mixed, Gypsy/Irish Traveller, Other White, Other, and Unknown. Where ethnicity was missing it was possible to supplement, for BiB children only, with data held by BiB (n=311).

To assess vulnerabilities in wellbeing, we grouped questions within four domains: (1) home, family and family relationships, (2) material resources, (3) friends and school, (4) subjective (self-reported) wellbeing. Within each domain, we defined ‘vulnerabilities’ on (a) the basis of research literature showing these to be well-established childhood risk factors for long-term health, wellbeing, educational attainment, and social mobility and (b) subsequent consultation with the vulnerabilities workstream of the Bradford Institute for Health Research Covid-19 Scientific Advisory Group (BIHR C-SAG) (https://www.bradfordresearch.nhs.uk/vulnerable-groups/). The risk factors categorised as vulnerabilities include, by domain:

- **Home, family, family relationships**: family never gets along well together, no garden where child can play, no nearby park where they can play, never plays in park
- **Material resources**: no warm winter coat, not having 3 meals a day, no internet at home, worried about money all the time
- **Friends and school**: doesn’t like school, not many friends, bullied some or all of the time, mean to others all the time, feel left out all the time
- **Subjective wellbeing**: never happy, always sad, ill/unwell all of the time, keeps worries to self, can never work out what to do when things are hard

### Data Analysis

Socio-demographics and the prevalence of vulnerabilities in child wellbeing are presented as counts and/or percentages with 95% confidence intervals, for all children and by gender and ethnicity. We also describe the number and percent of children with multiple vulnerabilities within and across domains of wellbeing by gender and ethnicity. Children with missing data for any answer are excluded from the relevant analyses and number of missing reported in figures and tables.

## Results

Table 1 shows the socio-demographics of the 15,641 survey respondents. Figure 1 presents the prevalence of each vulnerability across all domains for all children; Figure 2 presents the percentage of all children who have zero, one and more than one vulnerability within each domain.

**Table 1:** Socio-demographic characteristics of 15,641 Bradford primary school children.

|  | Number | Percent |
| --- | --- | --- |
| **Gender** |  |  |
| Female | 7647 | 48.9% |
| Male | 7994 | 51.1% |
| **Age (years)** |  |  |
| 6 | 2 | <0.01% |
| 7 | 5264 | 33.7% |
| 8 | 6901 | 44.1% |
| 9 | 3175 | 20.3% |
| 10 | 293 | 1.9% |
| 11 | 1 | <0.01% |
| 12 | 3 | <0.01% |
| 13 | 2 | <0.01% |
| Missing |  |  |
| **Ethnicity** |  |  |
| Pakistani | 7031 | 45.0% |
| Bangladeshi | 471 | 3.0% |
| Indian | 357 | 2.3% |
| Black/Black British | 277 | 1.8% |
| White British | 4247 | 27.1% |
| Mixed | 900 | 5.7% |
| Gypsy/Irish Traveller | 190 | 1.2% |
| White Other | 707 | 4.5% |
| Other | 425 | 2.7% |
| Unknown | 1036 | 6.6% |

**Figure 1:**
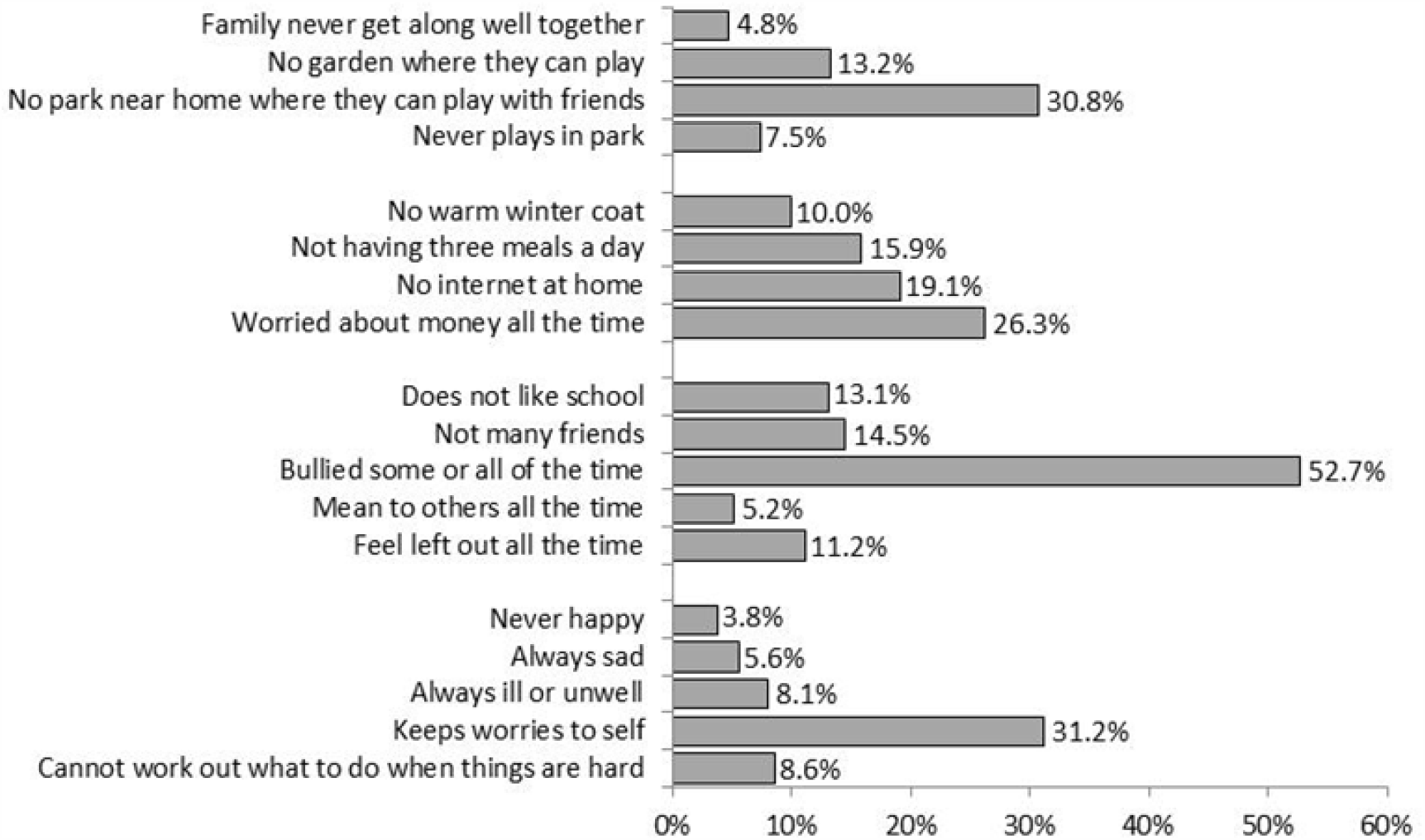
Percent of primary school children with each vulnerability across domains of wellbeing*. *For 15,641 children: each item has missing data of 3% or less, with the exception of items “No winter coat”, “Not having three meals a day” and “No internet at home” which have around 10% missing data. For more details, and confidence intervals, see Supplementary Materials: Table S1.

**Figure 2:**
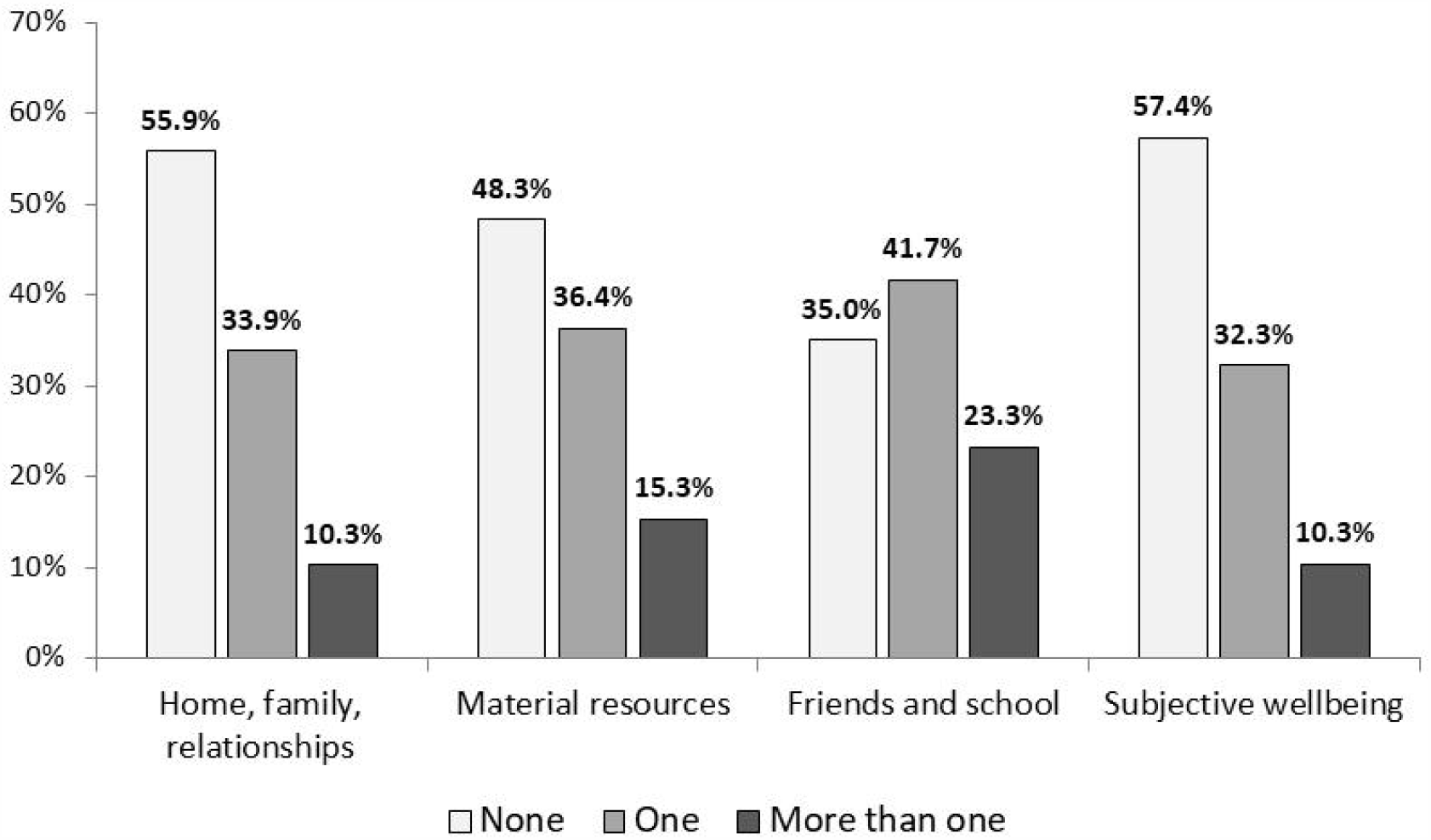
Percentage of primary school children with zero, one, or more than one vulnerability in each domain of wellbeing*. *For 15,641 children: the percentage with missing data for at least one item in each domain is between 5% and 16%. For more details, and confidence intervals, see Supplementary Materials: Table S2.

Although the prevalence of vulnerability varies by item and domain, within each domain there is an item with notably high prevalence, with more than one in four children reporting vulnerability: no park near home (30.8%, 95% CI: 30.1-31.5%); worried about money all the time (26.3%, 95% CI: 25.6-27.0%); bullied some or all of the time (52.7%, 95% CI: 51.9-53.4%); and keeping worries to self (31.2%, 95% CI: 30.5-31.9%). We considered six vulnerabilities in child wellbeing would be of particular concern during lockdowns and school closures associated with the Covid-19 pandemic: family never gets along well together; no garden where child can play; no nearby park where they can play; not having 3 meals a day; no internet at home; worried about money all the time. Before the pandemic 37.4% (36.6-38.3%) of children had one of these vulnerabilities and a further 29.6% (28.9-30.4%) had more than one.

Although the majority of children have no vulnerabilities in the Home, Family, Relationships domain (55.9%, 95% CI: 55.0-56.7%) and the Subjective Wellbeing domain (57.4%, 95% CI: 56.6-58.2%), the majority of children have one or more vulnerabilities in the Material Resources, and Friends and School domains. Only 10% (add 95% CI) of children have no vulnerabilities in any domain of wellbeing. Figure 3 is a Venn diagram that shows how vulnerabilities overlap across domains. 1,494 (10%) children have no vulnerabilities. Each oval in the Venn diagram includes all the children who have one or more vulnerabilities within that domain; so, for example, 2% of children have vulnerabilities in Home, Family, Relationships *and* Subjective Wellbeing but not in other domains. 1,519 children (10%) have one or more vulnerabilities in all four domains.

**Figure 3:**
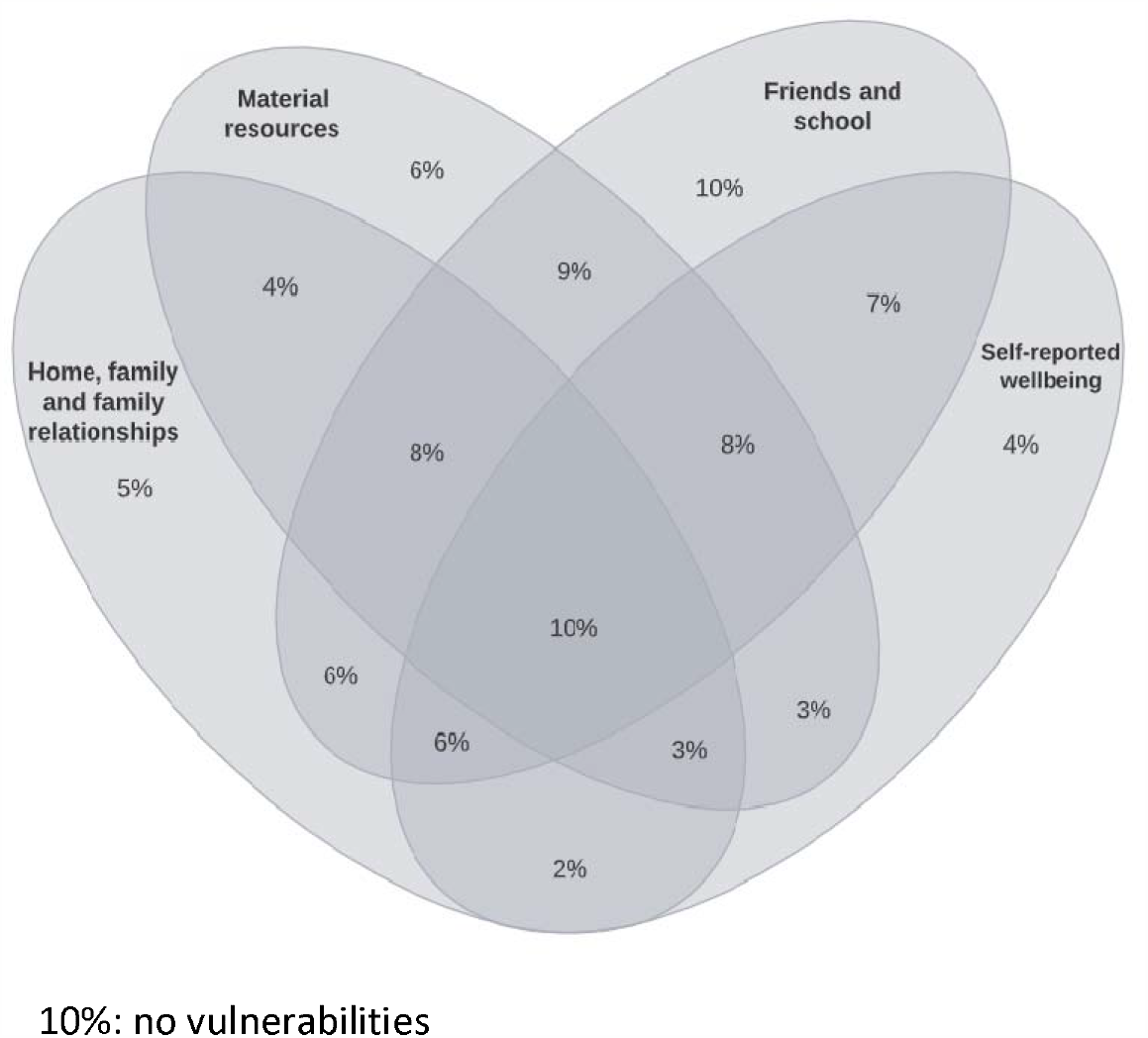
Percentage of children with vulnerabilities within and across domains. **(N=15,641)**

### Variation by gender

Boys were consistently significantly more likely to report all of the vulnerabilities in the Home, Family and Family Relationships domain, although the gender differences were not large (see Supplementary Materials: Table 3 for full results, including 95% confidence intervals, for all domains by gender). There were no gender differences in having access to the internet at home, but boys were significantly more likely to report not having a winter coat (11.1%, 95% CI: 10.4-11.9% vs 8.9%, 95% CI: 8.2-9.6%), not having three meals a day (17.7%, 95% CI: 16.8-18.6%) vs 14.0%, 95% CI: 13.3-14.9%) and worrying about money all the time (28.9%, 95% CI: 28.0-30.0%) vs. 23.4%, 95% CI: 22.5-24.4%). Within the Friends and School domain, boys were significantly more likely to not like school (18.0%, 95% CI: 17.1-18.8%) vs. 8%, 95% CI: 7.4-8.6%) and to be mean to others all the time (7.0%, 95% CI: 6.4-7.6% vs. 3.4%, 95% CI: 3.0-3.8%), while girls were significantly more likely to report not having many friends (16.7%, 95% CI: 15.9-17.6% vs. 12.5%, 95% CI: 11.8-13.2%), being bullied some or all of the time (55.8%, 95% CI: 54.7-56.9% vs. 49.7%, 95% CI: 48.6-50.8%) and feeling left out all the time (12.1%, 95% CI: 11.4-12.8%) vs. 10.3%, 95% CI: 9.7-11.0%). Within the Subjective Wellbeing domain there were no gender differences in being always sad or always ill or unwell, but boys were significantly likely to report never being happy (5.2%, 95% CI: 4.7-5.7%) vs. 2.4%, 95% CI: 2.1-2.8%), keeping worries to themselves (33.7%, 95% CI: 32.7-34.8% vs. 28.5%, 95% CI: 27.5-29.6%) and not being about to work out what to do when things are hard (9.6%, 95% CI: 9.0-10.3% vs. 7.5%, 95% CI: 6.9-8.1%). In all domains except for Friends and School, boys were significantly more likely to have at least one vulnerability, compared to girls.

### Variation by ethnicity

Figures 4a-4d shows the percentages (and 95% confidence intervals) of primary school children with at least one vulnerability within each of the domains of child wellbeing, by ethnicity. Full results are given in Supplementary Materials, Tables S4a-4d.

**Figure 4a:**
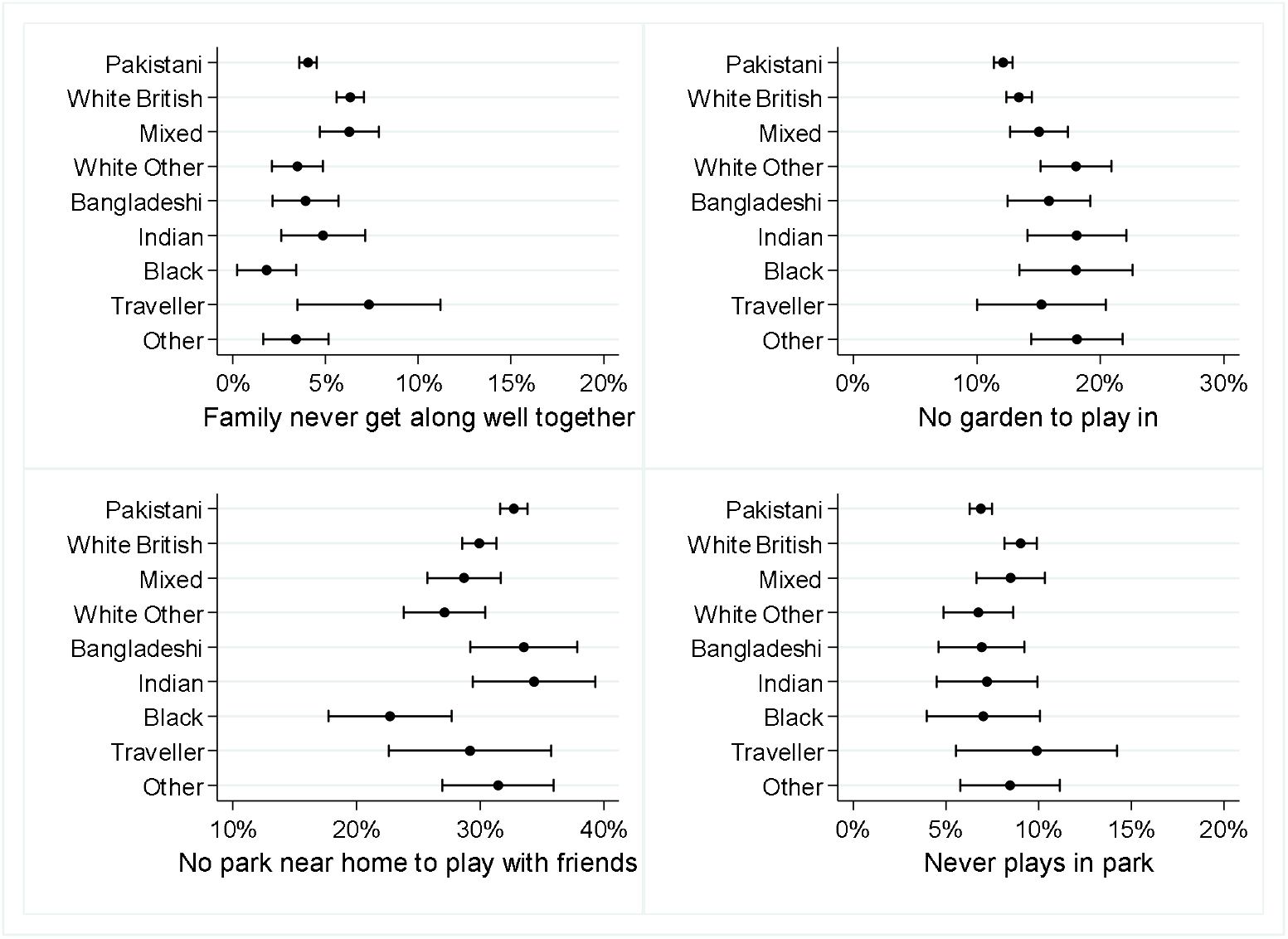
Percentage of primary school children with at least one vulnerability in the Home, Family and Family Relationships domain.

**Figure 4b:**
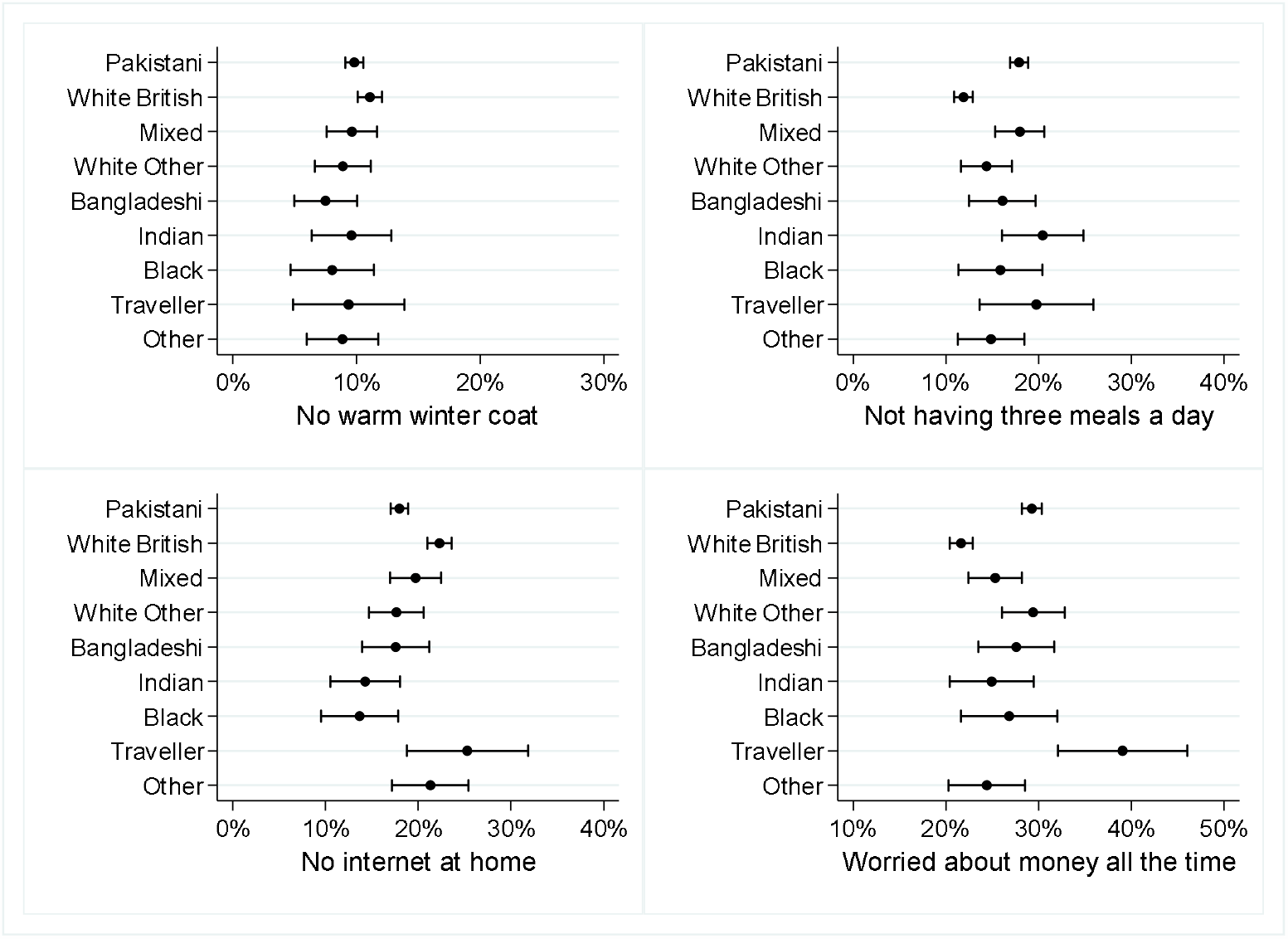
Percentage of primary school children with at least one vulnerability in the Material Resources domain.

**Figure 4c:**
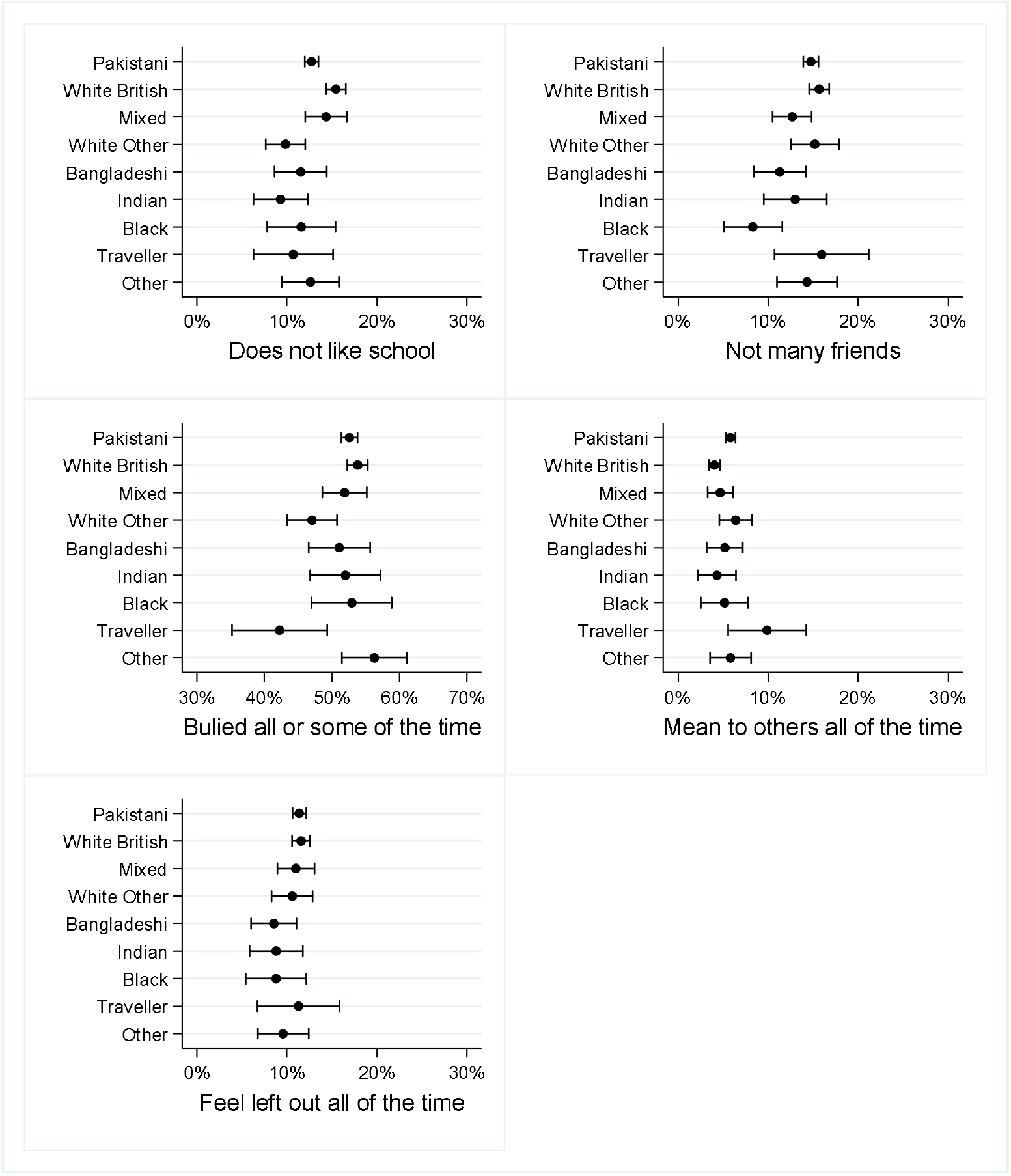
Percentage of primary school children with at least one vulnerability in the Friends and School domain.

**Figure 4d:**
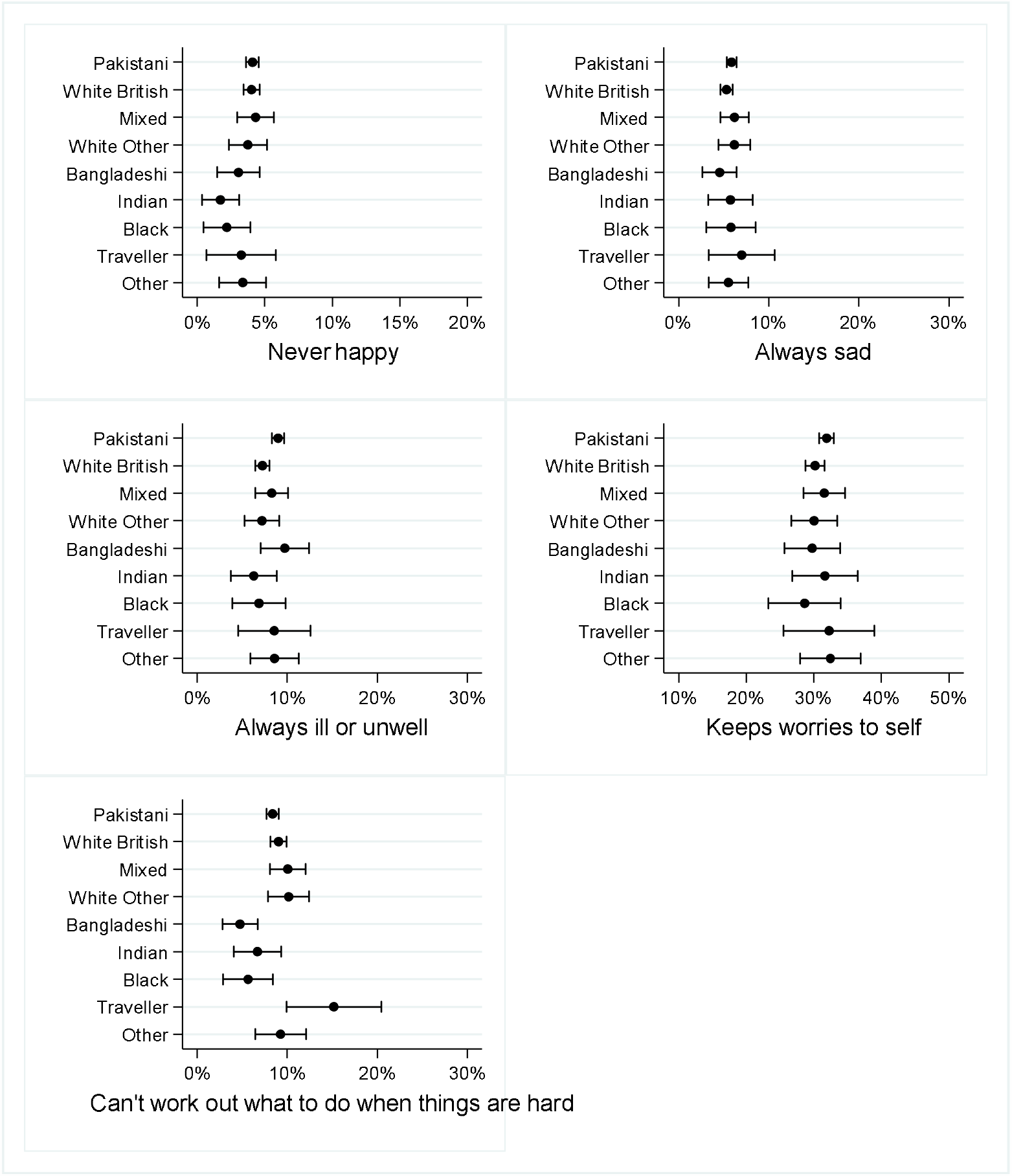
Percentage of primary school children with at least one vulnerability in the Subjective Wellbeing domain.

#### Home, family, relationships

Compared to White British children, Pakistani, White Other and Black/Black British children were significantly less likely to say their family never get along well together. White Other and Other children were significantly more likely to not have a garden where they can play, compared to White British children. Compared to White British children, Pakistani children were significantly more likely, and Black/Black British children significantly less likely to have no park near home where they can play with friends. Compared to White British children, Pakistani children were significantly less likely to say they never play in a park.

#### Material resources

There were no significant ethnic differences in the proportion of children who did not have a warm winter coat. Compared to White British children, Pakistani, Indian and Gypsy/Traveller children, as well as children of Mixed ethnicity were significantly more likely to report not having three meals a day. Pakistani, White Other, Indian and Black/Black British children were significantly less likely to report having no internet at home than White British children. Compared to White British children, Pakistani, White Other, Bangladeshi and Gypsy/Irish Traveller children were significantly more likely to worry all the time about how much money their families have.

#### Friends and school

Pakistani, White Other and Indian children were significantly less likely to report not liking school than White British children, and Bangladeshi and Black/British were significantly less likely to report not having many friends. Compared to White British children, White Other and Gypsy/Irish Traveller children were significantly less likely to say they were bullied some or all of the time, and Pakistani children and Gypsy/Irish traveller children were significantly more likely to say they were mean to others all the time. There were no significant ethnic differences in the proportion of children who felt left out by others all the time.

#### Subjective wellbeing

There were no significant ethnic differences in the proportion of children who said they were never happy or were always sad or who said they keep worries to themselves. Compared to White British children, only Pakistani children were significantly more likely to say they were always ill or unwell. Compared to White British children, Bangladeshi children were significantly less likely, and Gypsy/Irish traveller children more likely, to say they cannot work out what to do when things are hard.

## Discussion

### Statement of Principal Findings

Although most primary school children aged 7-10 in Bradford have good levels of wellbeing on most indicators across multiple domains, fewer than 10% have no vulnerabilities at all, and a worrying 10% have more than one vulnerability in all the four domains we studied. Variations in vulnerabilities by ethnicity were complex, with children in Black, Asian and Minority Ethnic groups sometimes reporting more vulnerabilities and sometimes fewer than White British children. We considered six vulnerabilities to be of particular concern during the current Covid-19 pandemic and associated national and local lockdowns (family never gets along well together; no garden where child can play; no nearby park where they can play; not having 3 meals a day; no internet at home; worried about money all the time). Before the pandemic, the majority – two-thirds – of Bradford children had one or more of these vulnerabilities. The domain with the highest proportion of children reporting vulnerabilities was Friends and School with 1 in 20 children reporting that they are mean to other children all the time, more than 1 in 10 reporting feeling left out all the time, not having many friends and not liking school, and the majority of children reporting being bullied some or all of the time.

We found that boys reported significantly more vulnerabilities for all indicators in the Home, Family and Family Relationships domain, as well as for the majority of indicators in the Material Resources, Friends and School, and Subjective Wellbeing domains. This contrasts with the finding of few consistent gender differences in wellbeing and a trend towards worse wellbeing in girls in the UK State of the Nation 2019: Children and Young People’s Wellbeing report^4^, and the general picture in the research literature of worse wellbeing for adolescent girls^15 16^ and inconsistent gender differences among adults.^17 18^ These differences may be due to the younger age of our sample, and suggest that worse wellbeing among adolescent girls might not be inevitable if prevention efforts were started before secondary education.

We found complex variations in vulnerabilities by ethnicity, with children in Black, Asian and Minority Ethnic groups sometimes reporting more vulnerabilities and sometimes fewer than White British children. For example, compared to children of Pakistani heritage, White British children were more likely to say that their family never gets along well and to have no access to the internet at home, whereas children with Pakistani heritage were more likely than White British children to say they had no park near their home where they can play with friends, to report not having three meals a day and to worry all the time about how much money their families have. The UK State of the Nation 2019: Children and Young People’s Wellbeing report drew attention to the importance of “*moving beyond the average*” in understanding wellbeing but reported “no discernible differences in wellbeing by…ethnicity” whilst recognising that small sample sizes might have obscured differences and reporting statistics only for 5 broadly defined ethnic groups. We provide a detailed description of wellbeing for 9 ethnic groups, so that nuances in ethnic variation can be better understood, including for smaller ethnic groups. For example, we found that Gypsy/Irish Traveller children were less likely than White British children to say they were bullied some or all of the time, but more likely to say they were mean to others, can never work out what to do when things are hard, worry all the time about how much money their family has, and not have 3 meals a day.

### Strengths and weaknesses of the study

To our knowledge, ours is the only large contemporary study of wellbeing in primary school aged children in England, with sufficient numbers of children belonging to some of the UK’s smaller Black, Asian and Minority Ethnic communities to be able to describe their wellbeing. Our approach to school recruitment, opt-out consent and whole classroom testing, along with Born in Bradford’s longstanding trusting relationships with schools and communities ensured a very high response rate. Although our study is based in a single city, it is likely to be generalizable to other UK cities with high levels of ethnic diversity and deprivation.

The fact that our measures of wellbeing are self-reported by children is both a strength and a potential weakness. It has been argued that, for adults and children alike, the subjective experience of quality of life matters, and that, as feelings have a demonstrable objective neuropsychological reality and are correlated with objective health, social and economic outcomes, they are worthy of measurement and inference^19 20^ Clearly, our measures of vulnerability not only include things that can only be measured subjectively, such as happiness or feeling left out, but also things that arguably would be better measured objectively, e.g., having access to a garden or three meals a day. However, it is possible that even for measures such as these, children’s perceptions that they lack material resources may be important even if they actually have the resources.

Our data collection objectives and approach to categorising vulnerabilities in wellbeing is very different to the Adverse Childhood Experiences (ACEs) approach^21^ and the definition and measurement of extreme child vulnerabilities collated and reported by the Children’s Commissioner for England (https://www.childrenscommissioner.gov.uk/chldrn/). ACEs are traumatic experiences that include abuse and neglect, parental mental illness, substance use and divorce. The child vulnerabilities reported by the Children’s Commissioner include living in a household where an adult has a drug or alcohol dependence, where an adult has experienced violence or abuse from a partner, where an adult has a clinically diagnosable mental health condition or more than one of these factors. Both ACEs and the Children’s Commissioner approaches identify factors that put children at immediate, as well as long-term, risk of harm. Our approach was to collect data on more common risk factors with known long-term consequences for health and wellbeing, to better understand children’s subjective experiences across the whole population.

In our school-based whole classroom surveys, time constraints and study design prevented data collection and analyses that would have allowed us to go beyond description and investigate the causes of vulnerabilities in child wellbeing. For BiB children only, we examined associations between neighbourhood area deprivation (Index of Multiple Deprivation) and vulnerabilities. We found no significant associations for any domain except for more vulnerabilities in Material Resources at higher levels of deprivation, and as lack of material resources is definitional for deprivation do not report those analyses here. We plan future data collection with this sample to allow us to examine how vulnerabilities at primary school ages shape trajectories of wellbeing across time.

### Implications for providers of services to children and policymakers

Our findings suggest that, prior to the Covid-19 pandemic, a substantial number of children were experiencing sub-optimal wellbeing with potential lifelong consequences, with implications for all providers of services to children, as well as local and national policymakers. As the economic impact of the pandemic is increasing child poverty, income insecurity, food and housing insecurity, parental mental health challenges and family violence, the need for attention to the identification and amelioration of vulnerability is heightened.^22^ The International Society for Social Pediatrics and Child Health has called for the adoption of “a Child Rights Based Approach (CRBA) to respond to the COVID-19 pandemic, and to advance a future in which the health, development and well-being of children and youth are prioritised with explicit strategies to reduce health inequities and advance social justice” (https://www.issop.org/2020/06/01/issop-covid-19-declaration/). Such an approach can underpin and inform services and policy making at all levels.

As the population was encouraged to ‘lockdown’ at home and schools closed for almost all children (and attendance was very low for those who were eligible to go to school) for varying periods, the response to the pandemic will have affected children’s wellbeing (as well as their educational trajectories) differently, depending on their circumstances and context. Children who found school a nurturing refuge from difficult home situations will have suffered from school shutdowns, whereas others who found school a challenging environment may have been happy to be at home for a prolonged period; most children have probably experienced both positive and negative impacts. Although all schools work hard to create safe and supportive environments for children, the high levels of bullying and social isolation that we report suggests that more needs to be done to make this a reality for all children. In addition, the high prevalence of children with no access to the internet at home and no access to a garden or park mean that many children will have at times been unable to study or be physically active.

Locally, our research team has engaged with our local authority, public health, NHS and voluntary sector colleagues to identify principles to mitigate the impact of lockdown exit on vulnerable groups (https://www.bradfordresearch.nhs.uk/wp-content/uploads/2020/04/CSAG-Briefing-Paper-Vulnerable-Groups-and-Recovery-230420.pdf). We have proposed additional support for children and young people in the lockdown exit and recovery planning, including assessing the impact of decision making on children, prioritising children’s needs, providing extra support for vulnerable children, and monitoring impacts on children’s lives and wellbeing.

## Conclusions and implications for public health

Among primary school children aged 7-10 in a deprived Northern city in the UK, fewer than 10% have no vulnerabilities in home, family and family relationships, material resources, friends and school and subjective wellbeing, while 10% have more than one vulnerability in all of these domains. These children are at risk of poor health, social and economic trajectories throughout their lives that could be mitigated by policy focused at structural factors, such as poverty eradication, and child services targeted at reducing inequalities and supporting the most vulnerable.

## Data Availability

Scientists are encouraged and able to use BiB data, which are available through a system of managed open access.

## Ethics approval

Ethical approval has been obtained from the National Health Service Health Research Authority Yorkshire and the Humber (Bradford Leeds) Research Ethics Committee (reference: 16/YH/0062).

## Transparency statement

Kate Pickett (the manuscript’s guarantor) affirms that the manuscript is an honest, accurate, and transparent account of the study being reported; that no important aspects of the study have been omitted; and that any discrepancies from the study as originally planned have been explained.

## Acknowledgements

This study was funded by a joint grant from the UK Medical Research Council and UK Economic and Social Science Research Council (MR/N024397/1), with support from a Wellcome Trust infrastructure grant (WT101597MA) and the National Institute for Health Research under its Applied Research Collaboration Yorkshire and Humber (NIHR200166). The funders had no role in the design of the study, the collection, analysis, or interpretation of the data; the writing of the manuscript, or the decision to submit the manuscript for publication. All authors are independent of the funders, had full access to all of the data (including statistical reports and tables) in the study and can take responsibility for the integrity of the data and the accuracy of the data analysis. Views expressed in this paper are those of the authors and not necessarily those of any funder.

Born in Bradford is only possible because of the enthusiasm and commitment of the Children and Parents in BiB. We are grateful to all the participants, health professionals, schools, and researchers who have made Born in Bradford happen.

### Summary

#### What is already known on this topic

- Child wellbeing in the UK is low in comparison to other rich countries
- Little is known about the wellbeing of children under the age of 10 in the UK
- Little is known about ethnic variations in child wellbeing in the UK

#### What this study adds

- Although most 7-10 year old children in our study have good levels of wellbeing, fewer than 10% have no vulnerabilities at all, a worrying 10% have at least one vulnerability in all the domains we studied, and two thirds have vulnerabilities of concern during the Covid-19 lockdowns
- In contrast to studies of older children, boys had more wellbeing vulnerabilities, although girls were more likely to have no friends, report being bullied and feeling left out.
- Ethnic variations in vulnerabilities in wellbeing were complex, with children from ethnic minorities reporting fewer of some vulnerabilities than White British children and more of others; our study is the first to report a wide range of vulnerabilities across a large number of ethnic groups in this age range

