## Supplementary materials for "Vulnerabilities in child wellbeing among primary school children: a cross-sectional study in Bradford, UK"

1: STROBE Statement—Checklist of items that should be included in reports of ***cross-sectional studies***

|  | **Item No** | **Recommendation** |
| --- | --- | --- |
| **Title and abstract** | 1 | (*a*) Indicate the study’s design with a commonly used term in the title or the abstract SEE TITLE |
|  |  | (*b*) Provide in the abstract an informative and balanced summary of what was done and what was found SEE ABSTRACT |
| **Introduction** | | |
| Background/rationale | 2 | Explain the scientific background and rationale for the investigation being reported  SEE INTRODUCTION, PAGE 4 |
| Objectives | 3 | State specific objectives, including any prespecified hypotheses  N/A |
| **Methods** | | |
| Study design | 4 | Present key elements of study design early in the paper  SEE METHODS, PAGES -7 |
| Setting | 5 | Describe the setting, locations, and relevant dates, including periods of recruitment, exposure, follow-up, and data collection  SEE SETTINGS AND PARTICIPANTS, PAGE 5 |
| Participants | 6 | (*a*) Give the eligibility criteria, and the sources and methods of selection of participants  SEE SETTINGS AND PARTICIPANTS, PAGE 5 |
| Variables | 7 | Clearly define all outcomes, exposures, predictors, potential confounders, and effect modifiers. Give diagnostic criteria, if applicable  SEE DESIGN AND PROCEDURES, PAGE 6 |
| Data sources/ measurement | 8* | For each variable of interest, give sources of data and details of methods of assessment (measurement). Describe comparability of assessment methods if there is more than one group  SEE DESIGN AND PROCEDURES, PAGE 6 |
| Bias | 9 | Describe any efforts to address potential sources of bias  SEE FIGURE 1, PAGE 5 |
| Study size | 10 | Explain how the study size was arrived at  SEE SETTINGS AND PARTICIPANTS, PAGE 5 |
| Quantitative variables | 11 | Explain how quantitative variables were handled in the analyses. If applicable, describe which groupings were chosen and why  SEE DESIGN AND PROCEDURES, PAGE 6 |
| Statistical methods | 12 | (*a*) Describe all statistical methods, including those used to control for confounding |
|  |  | (*b*) Describe any methods used to examine subgroups and interactions |
|  |  | (*c*) Explain how missing data were addressed |
|  |  | (*d*) If applicable, describe analytical methods taking account of sampling strategy |
|  |  | (*e*) Describe any sensitivity analyses  SEE DATA ANALYSIS, PAGE 7 |
| **Results** | | |
| Participants | 13* | (a) Report numbers of individuals at each stage of study—eg numbers potentially eligible, examined for eligibility, confirmed eligible, included in the study, completing follow-up, and analysed |
|  |  | (b) Give reasons for non-participation at each stage |
|  |  | (c) Consider use of a flow diagram  SEE METHODS, FIGURE 1, PAGE 5 AND ALL RESULTS TABLES |
| Descriptive data | 14* | (a) Give characteristics of study participants (eg demographic, clinical, social) and information on exposures and potential confounders   SEE TABLE 1, PAGE 8 |
|  |  | (b) Indicate number of participants with missing data for each variable of interest  SEE ALL TABLES AND FIGURES |
| Outcome data | 15* | Report numbers of outcome events or summary measures  SEE FIGURE 1, PAGE 9 |
| Main results | 16 | (*a*) Give unadjusted estimates and, if applicable, confounder-adjusted estimates and their precision (eg, 95% confidence interval). Make clear which confounders were adjusted for and why they were included  SEE ALL TABLES AND FIGURES |
|  |  | (*b*) Report category boundaries when continuous variables were categorized  N/A |
|  |  | (*c*) If relevant, consider translating estimates of relative risk into absolute risk for a meaningful time period  N/A |
| Other analyses | 17 | Report other analyses done—eg analyses of subgroups and interactions, and sensitivity analyses  SEE DISCUSSION RE: UNREPORTED ANALYSIS OF IMD, PAGE 18 |
| **Discussion** | | |
| Key results | 18 | Summarise key results with reference to study objectives  SEE STATEMENT OF PRINCIPAL FINDINGS, PAGE 16 |
| Limitations | 19 | Discuss limitations of the study, taking into account sources of potential bias or imprecision. Discuss both direction and magnitude of any potential bias  SEE STRENGTHS AND WEAKNESSES OF THE STUDY, PAGE 17 |
| Interpretation | 20 | Give a cautious overall interpretation of results considering objectives, limitations, multiplicity of analyses, results from similar studies, and other relevant evidence  SEE IMPLICATIONS SECTION, PAGE 18 |
| Generalisability | 21 | Discuss the generalisability (external validity) of the study results  SEE STRENGTHS AND WEAKNESSES OF THE STUDY, PAGE 17 |
| **Other information** | | |
| Funding | 22 | Give the source of funding and the role of the funders for the present study and, if applicable, for the original study on which the present article is based  SEE FUNDING STATEMENT |

**2. Me and My Life wellbeing survey**

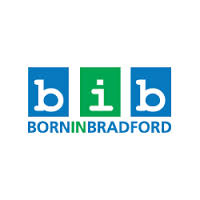

**Me and my life**

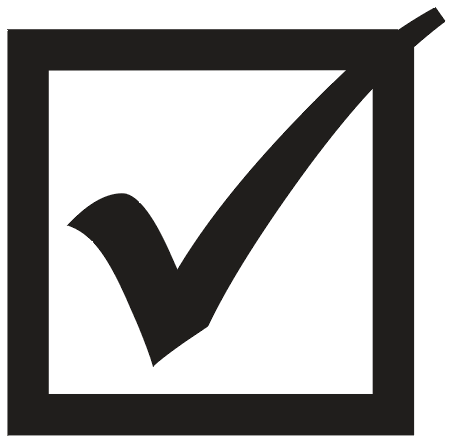

Please tick the box next to your answer

Please try to answer all the questions. There are no right or wrong answers. We want to know what you think.

Your answers will not be shown to anyone that you know (including mums and dads).

If you do not want to answer a question you can miss it out.

If you need any help, you can ask your teacher or the person who gave you this quiz.

**Thank you!**

1. Which of these would you most like to eat today?

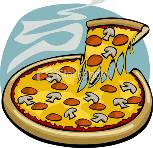

Pizza

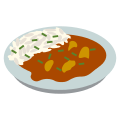

Curry

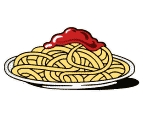

Pasta with sauce

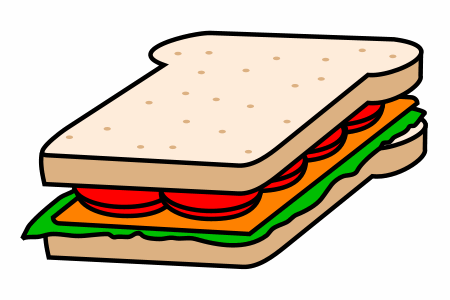

Sandwich

**Where I live**

2. How much do you like the home where you live?

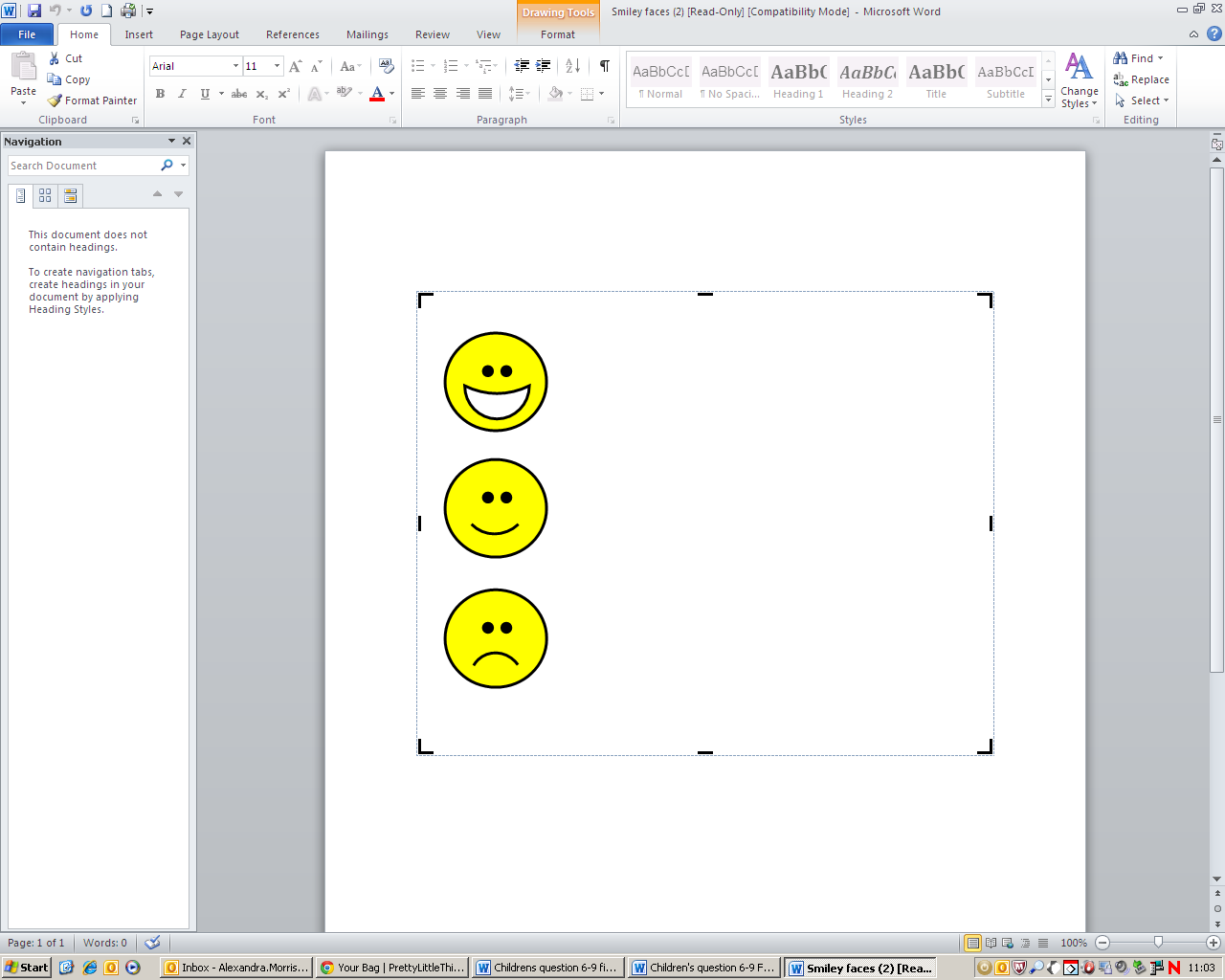

I like it a lot

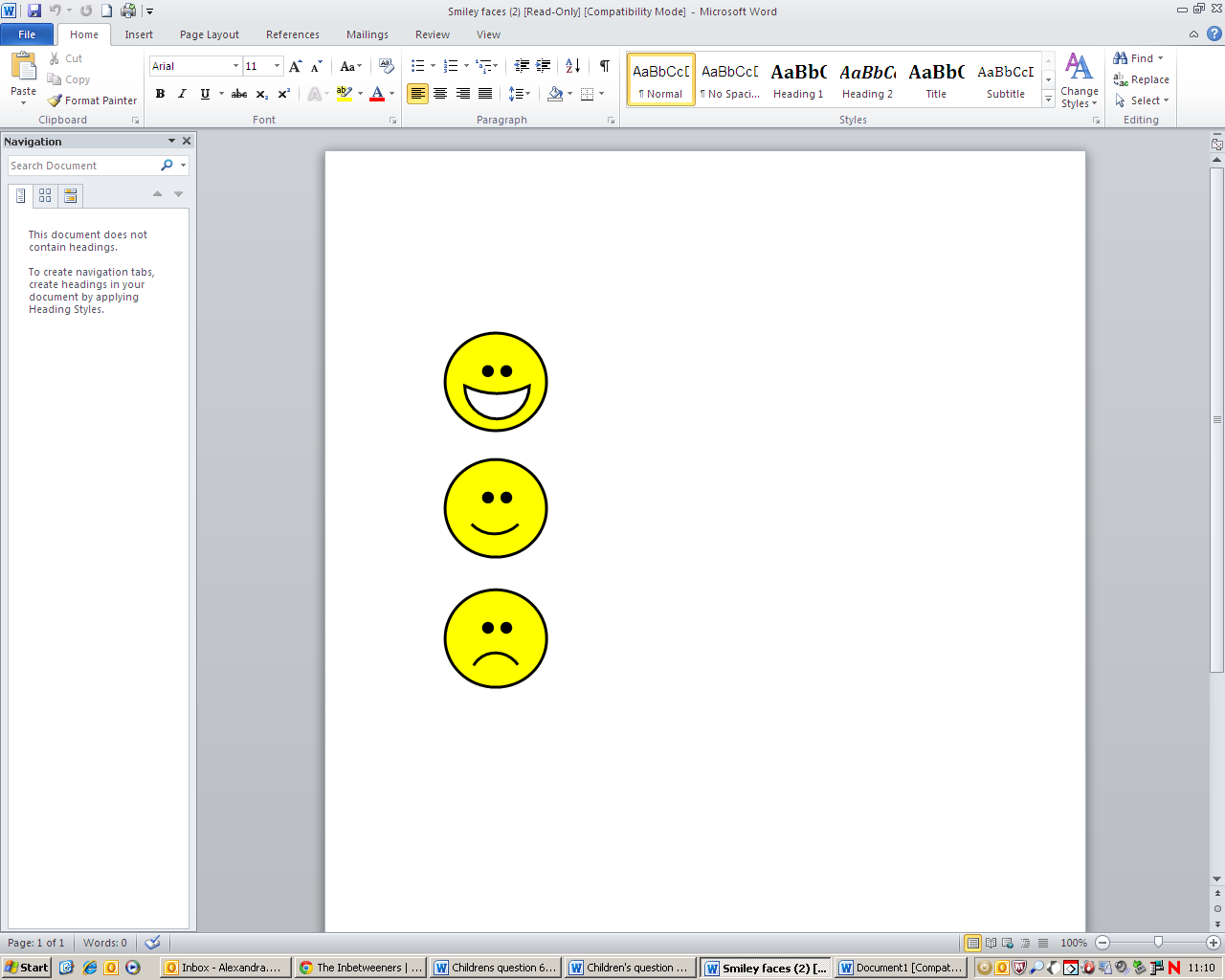

I like it a bit

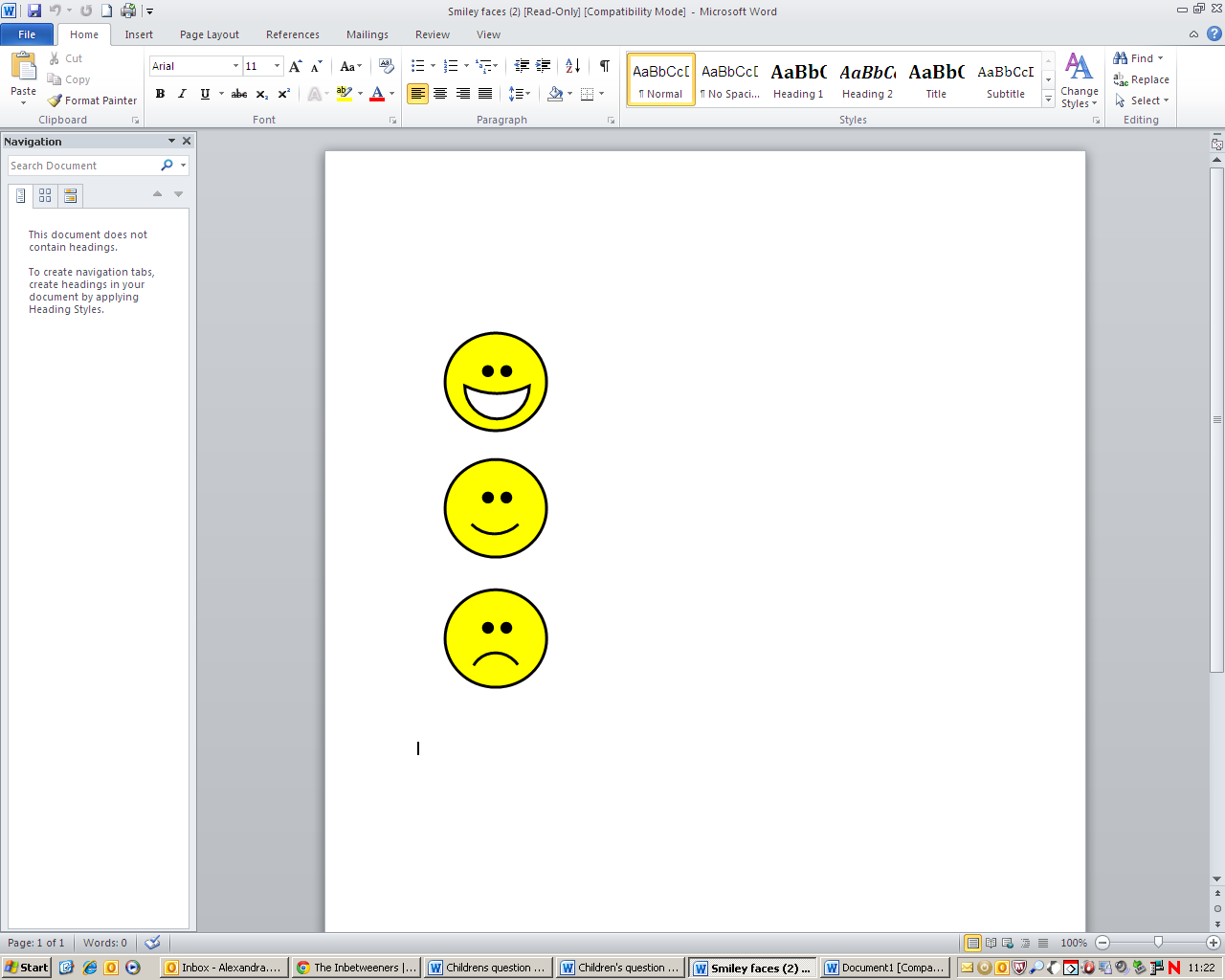

I don’t like it

3. How often do you play in a park?

Very often

Sometimes

Never

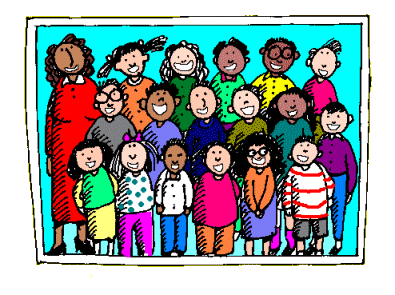
**My family and the people I live with**

4. How often do you have fun with your family at the weekend?

All of the time

Some of the time

Never

5. How often does your family get along well together?

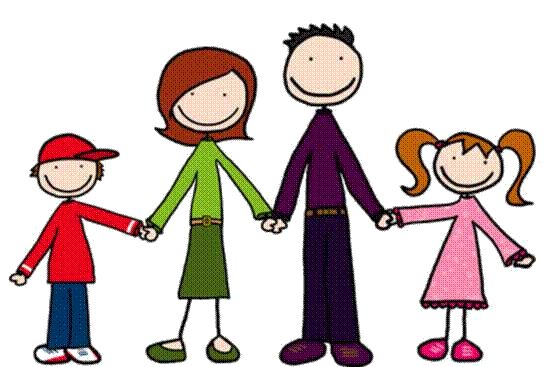

All of the time

Some of the time

Never

6. How often do you get along with your brothers, sisters and other children you live with?

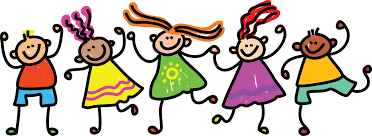
All of the time

Some of the time

Never

I don’t have brothers or sisters or live with other children

7. What languages do you speak with your family at home?

You can tick **more than one** answer if you like.

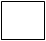

English

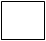

Urdu

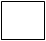

Mirpuri/Punjabi

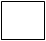

Slovakian

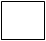
Arabic

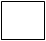

Bangla/Bengali

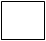

Hindko

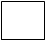
Pushto

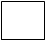

Other

**A bit about me**

8. Here is a list of things that some children have.

Do you have each of these things?

|  | **Yes**  **I have it** | **No**  **I don’t have it** |
| --- | --- | --- |
| 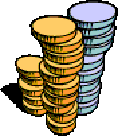  **Some money to spend on myself** | 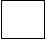 | 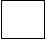 |
| 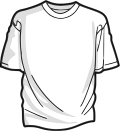  **Clothes that you think your friends like** | 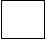 | 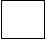 |
| 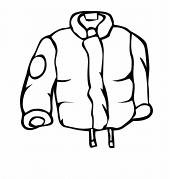  **A warm winter coat** | 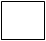 | 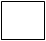 |
| 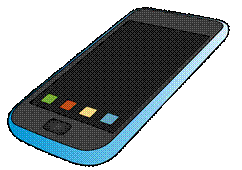  **Your own mobile phone** | 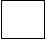 | 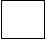 |
| 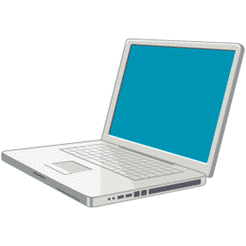  **A computer, laptop or tablet with internet at home** | 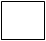 | 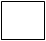 |
| 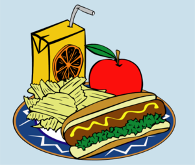  **Three meals every day** | 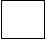 | 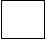 |

9. Do you have each of these things?

|  | **Yes**  **I have it** | **No**  **I don’t have it** |
| --- | --- | --- |
| 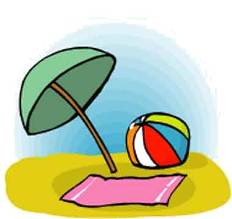  **Family trips or days out at least once a month, such as to the seaside or a theme park** | 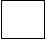 | 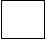 |
| 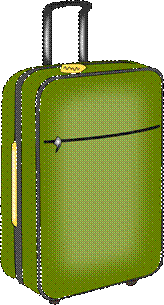  **At least one family holiday away from home each year** | 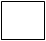 | 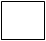 |

10. Does your home have a garden where you can play?

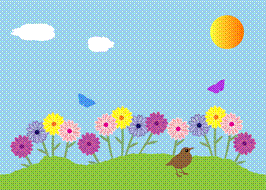

Yes

No

11.
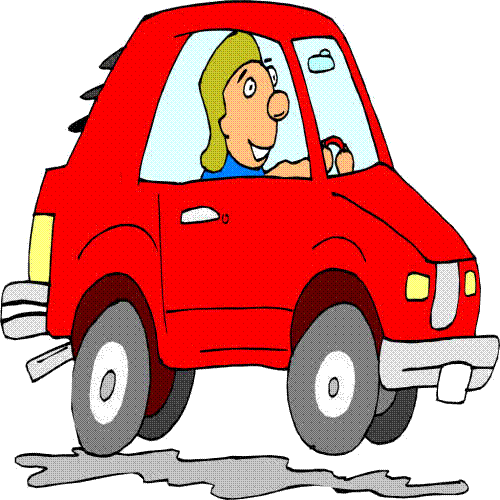
Does your family have a car?

Yes

No

12. Do you have a park near your home where you can play with your friends?

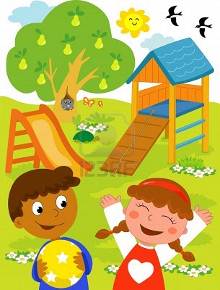

Yes

No

13. How often do you worry about how much money your family has?

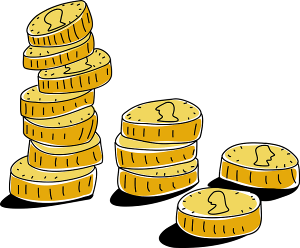

All of the time

Some of the time

Never

14.
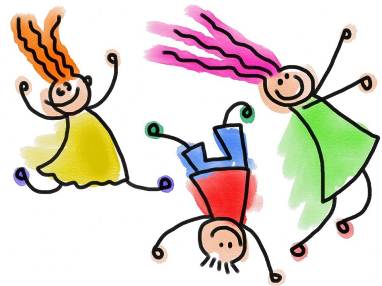
How often do you feel happy?

All of the time

Some of the time

Never

15. How often do you feel sad?

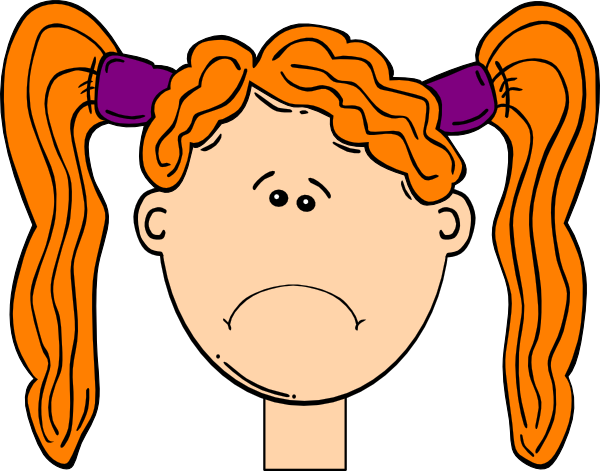

All of the time

Some of the time

Never

16.
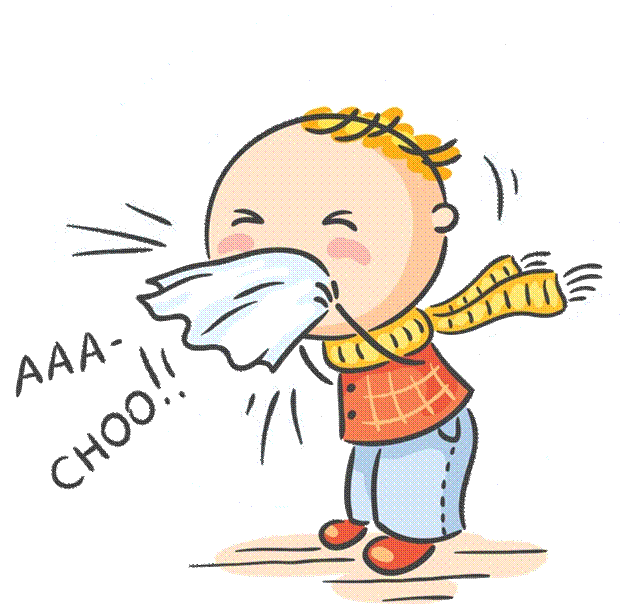
How often are you ill or unwell?

All of the time

Some of the time

Never

17. How often do you feel healthy?

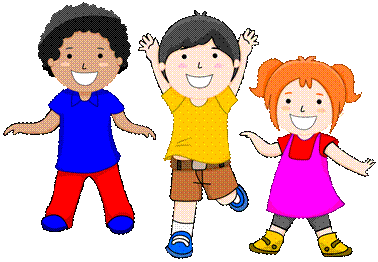

All of the time

Some of the time

Never

18. What do you do if you are worried about something?

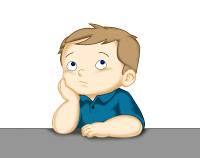

You can tick **more than one** answer if you like.

I keep it to myself

I tell a friend

I tell my Mum, Dad or the

person who looks after me

I tell a teacher

19. When I find something really hard I can work out what to do next.

All of the time

Some of the time

Never

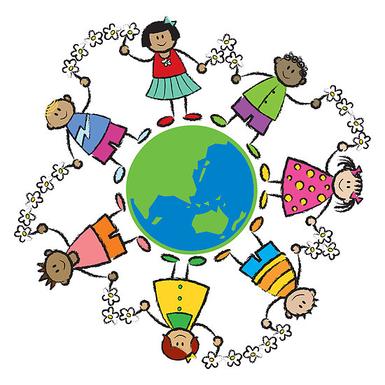
**My friends and other children**

20. How many friends do you have?

Lots

Some

Not many

21. Do you have any best friends?

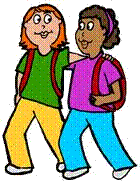

Yes

No

22. How much do you like playing with your friends?

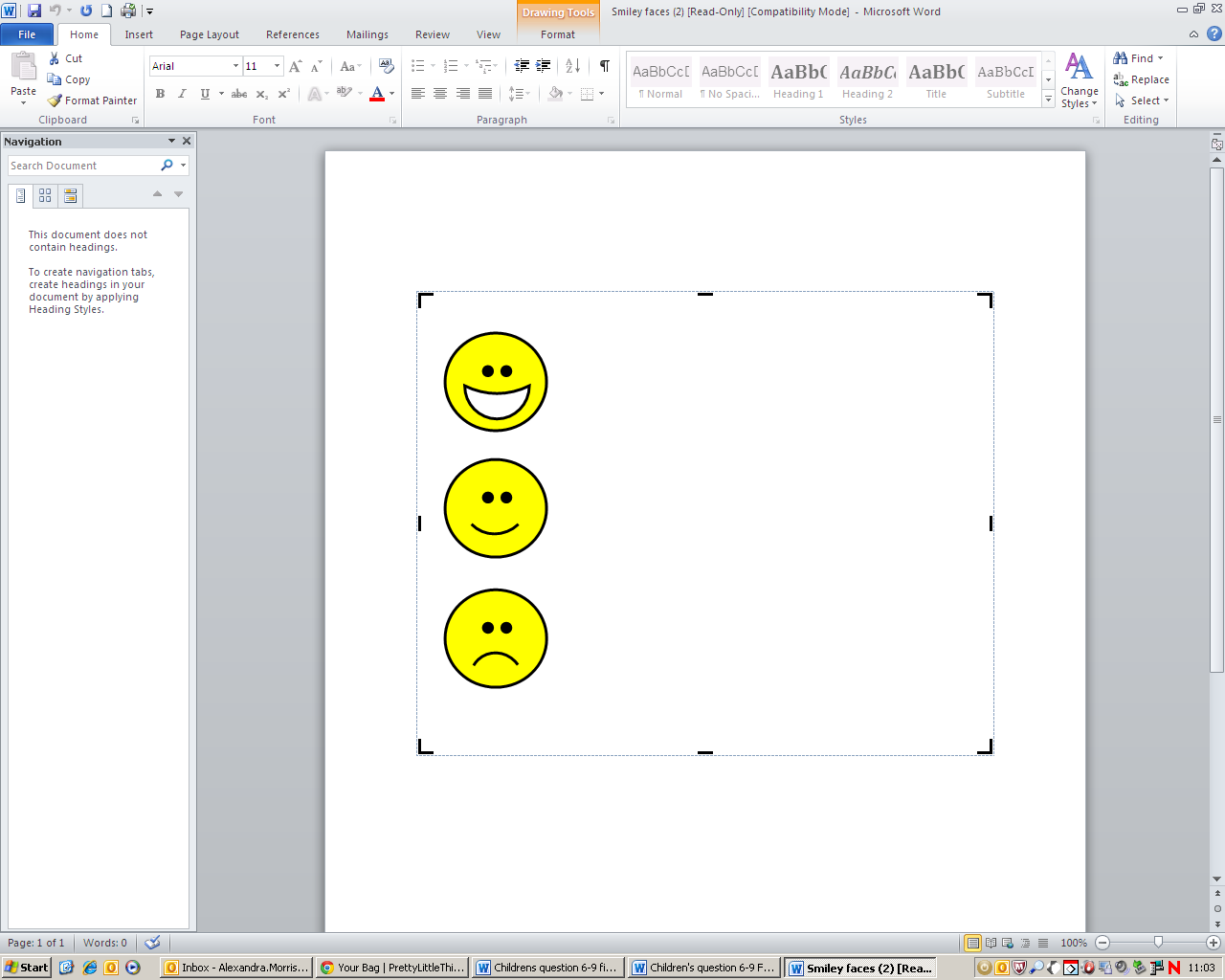

I like it a lot

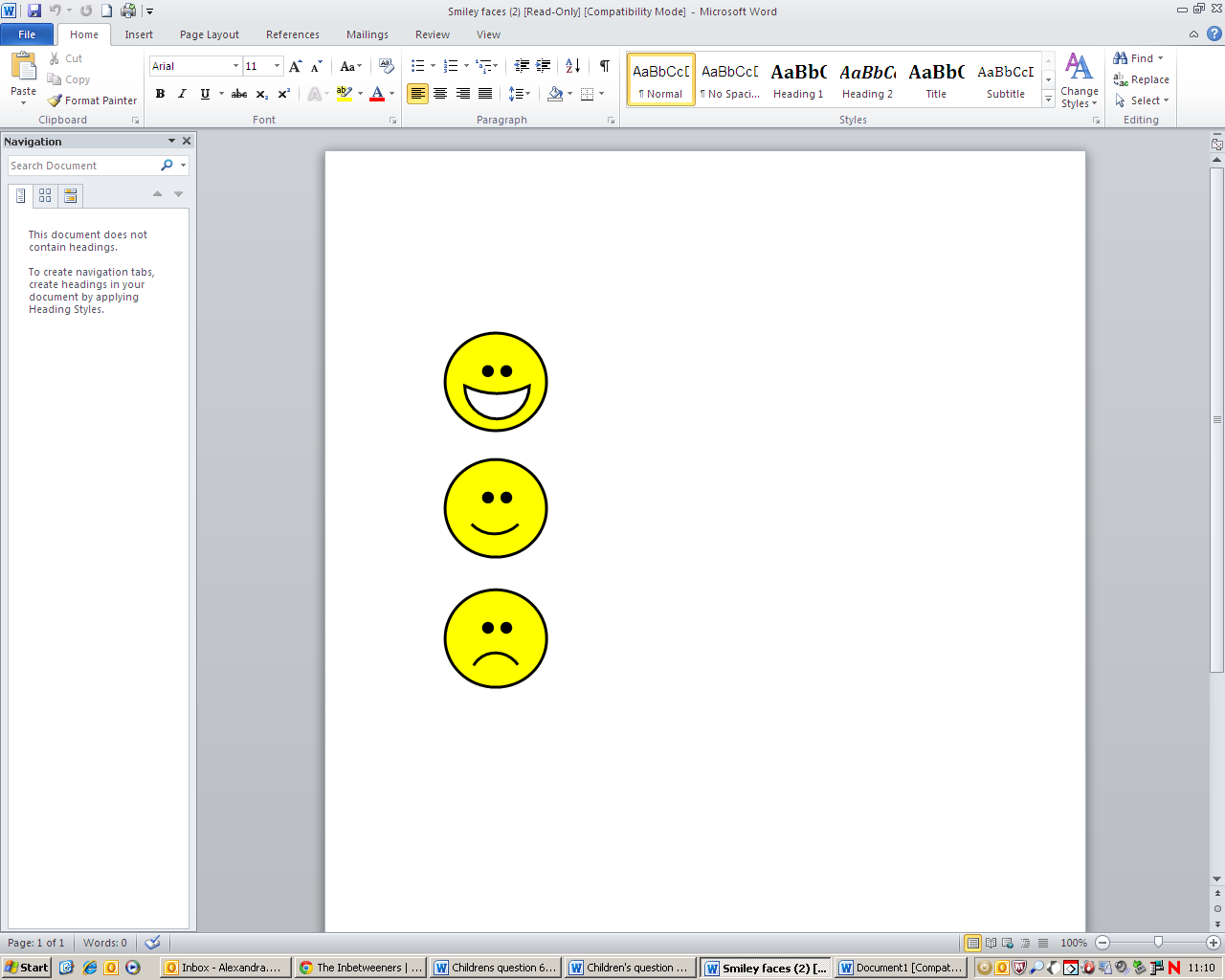

I like it a bit

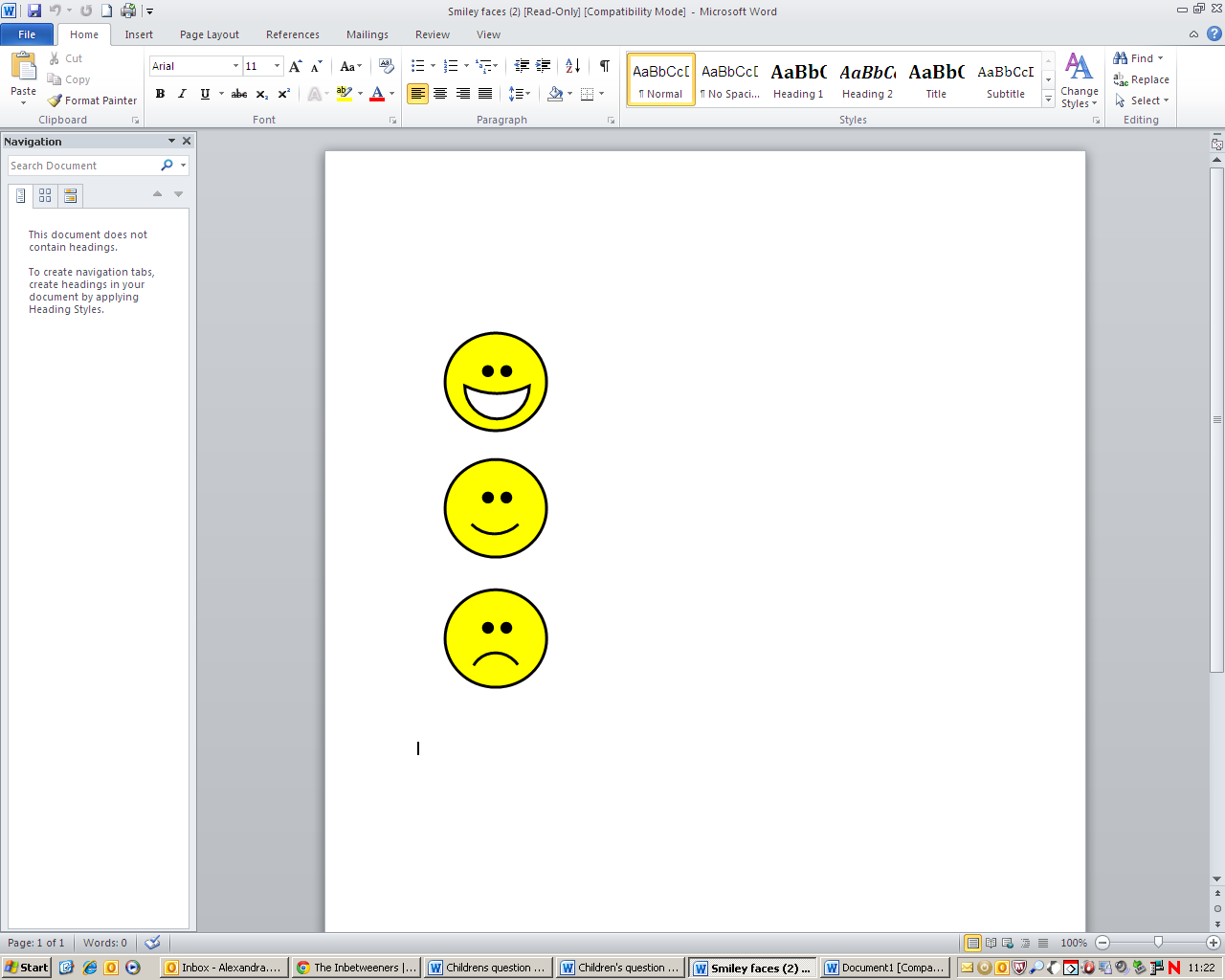

I don’t like it

23. How often do other children bully you?

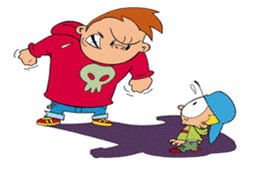
All of the time

Some of the time

Never

24. How often are you mean to other children at school?

All of the time

Some of the time

Never

25. How often do you feel left out of things by other children?

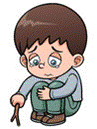

All of the time

Some of the time

Never

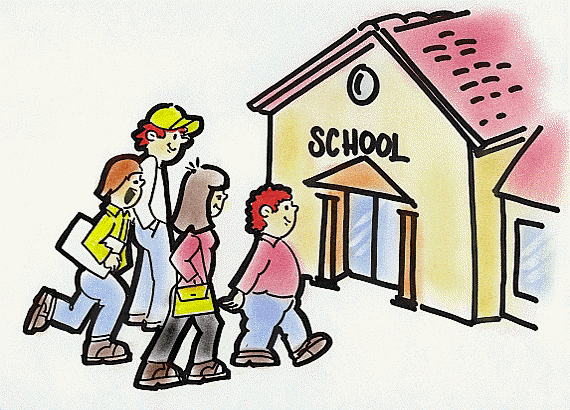
**My school**

26. How much do you like school?

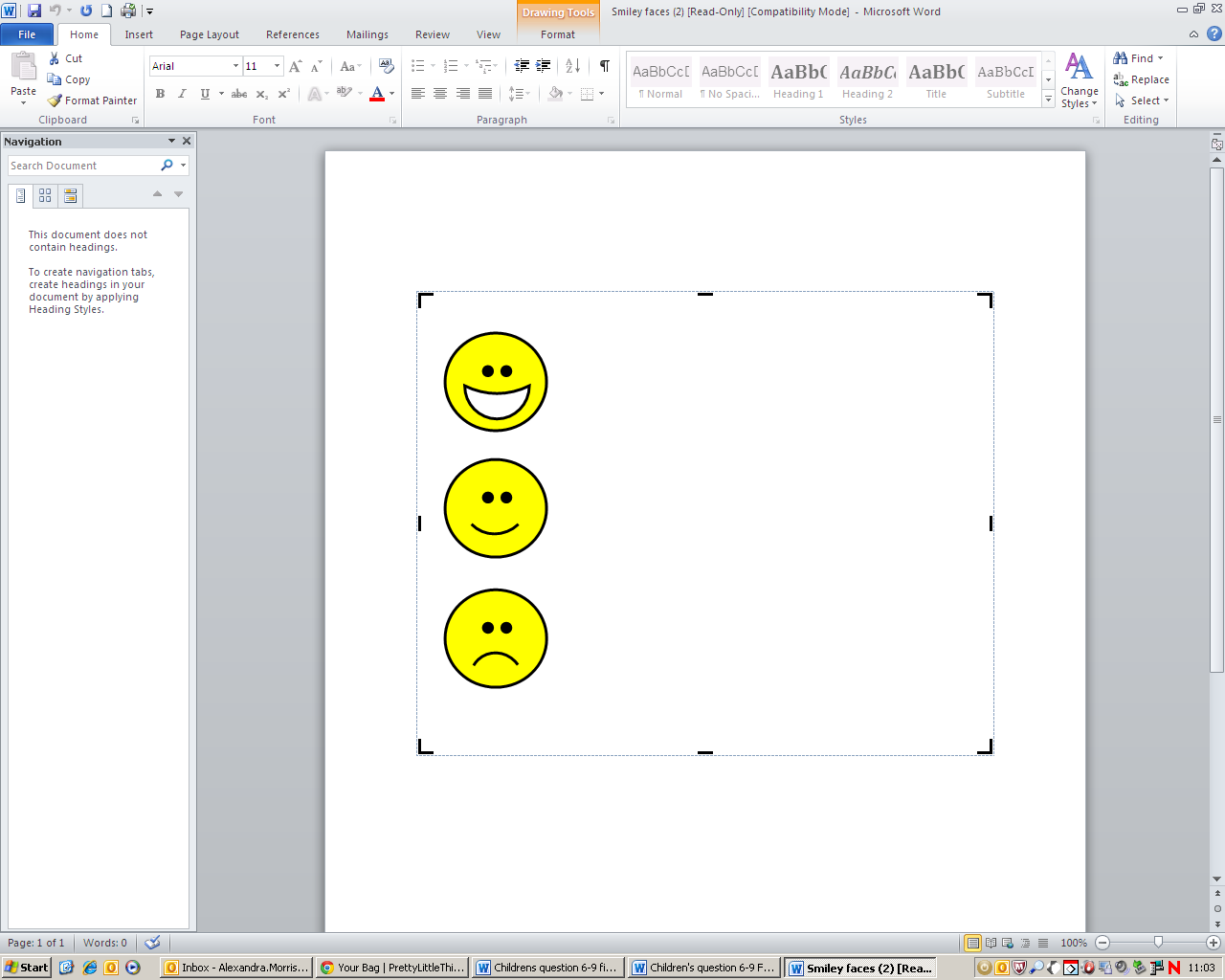

I like it a lot

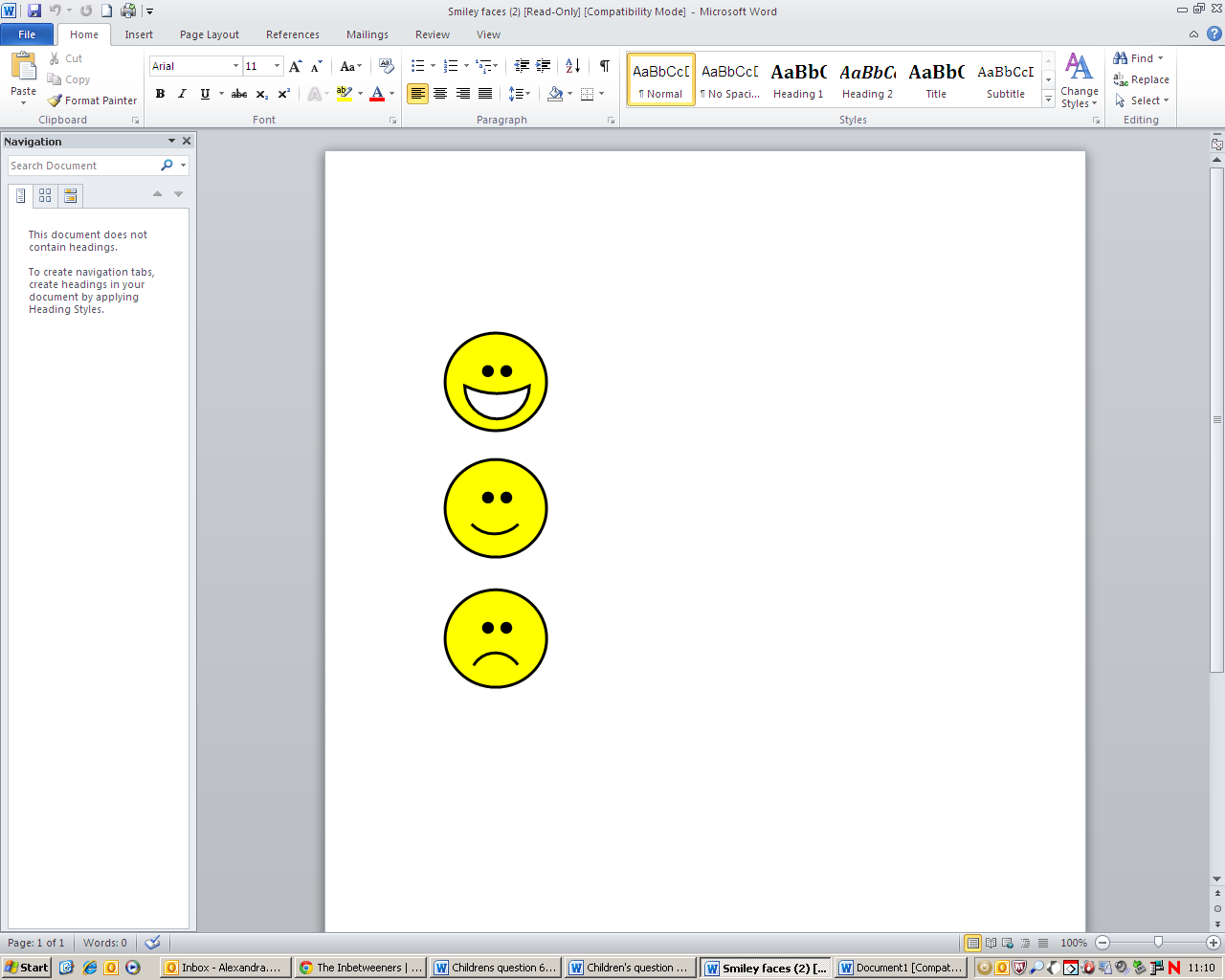

I like it a bit

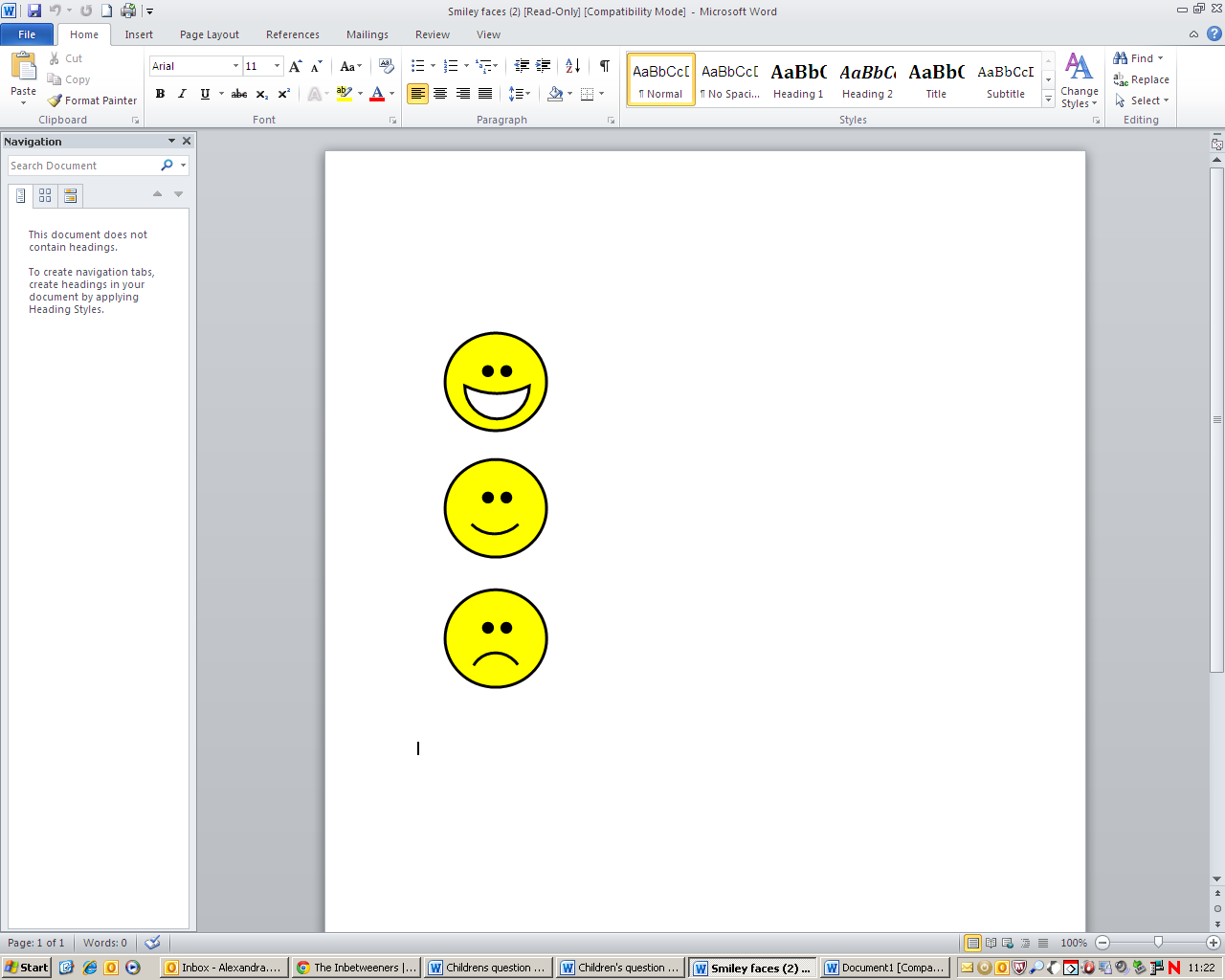

I don’t like it

**After school and weekend activities**

27. Do you go to any of these clubs or activities after school or at weekends?

|  | **Yes** | **No** |
| --- | --- | --- |
| 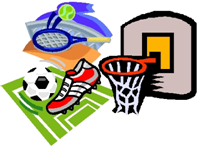  **Sports**  **(such as football, cricket, swimming, karate)** | 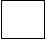 | 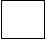 |
| 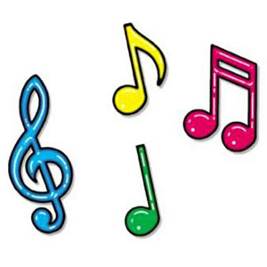  **Music lessons**  **(such as choir, piano lessons)** | 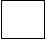 | 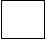 |
| 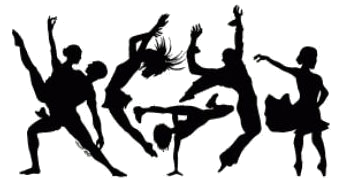  **Dance lessons** | 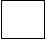 | 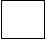 |
| **Any other club**  **(such as art, drama, film, cooking)** | 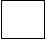 | 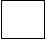 |

**Religion**

28. **What is your religion?**

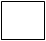

Christian

Muslim

Hindu

Sikh

Other religion

None/No religion

Don’t know

**If you are Muslim:**

**Do you go to Mosque or Madrassa?**

Yes

No

**If yes: How often do you go to Mosque or Madrassa?**

Most days a week

Some days a week

Less than once a week

**If you are Christian:**

**Do you go to Church or Sunday school?**

Yes

No

**If yes: How often do you go to Church or Sunday school?**

Most days a week

Some days a week

Less than once a week

**If you are Hindu/Sikh:**

**Do you go to a Temple?**

Yes

No

**If yes: How often do you go to a Temple?**

Most days a week

Some days a week

Less than once a week

**If you are another religion:**

**Do you go to a place of worship?**

Yes

No

**If yes: How often do you go to a place of worship?**

Most days a week

Some days a week

Less than once a week

**When I grow up**

29. When you grow up, would you like to be:

|  | **Yes, I’d like it a lot** | **Yes, I’d like it a bit** | **No, not at all** |
| --- | --- | --- | --- |
| 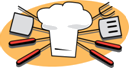  **A cook** | 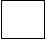 | 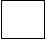 | 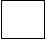 |
| 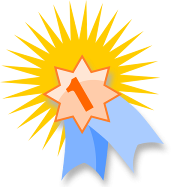  **A sports star** | 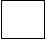 | 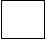 | 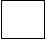 |
| 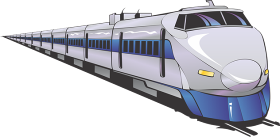  **A train driver** | 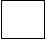 | 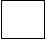 | 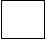 |
| 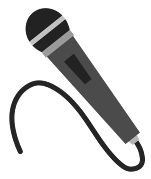  **A pop star** | 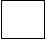 | 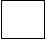 | 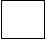 |
| 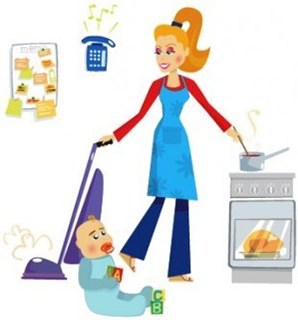  **Someone who looks after children at home** | 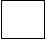 | 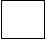 | 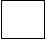 |
| 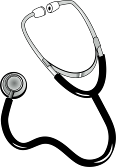  **A doctor or a nurse** | 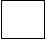 | 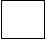 |  |
| **Someone who works in a shop** |  |  |  |
| **A teacher** |  |  |  |
| **Someone who works with computers** |  |  |  |

30. What would you most like to be when you grow up?

You can write any job you like!

………………………………………………………………………………………………………………………………………………..

………………………………………………………………………………………………………………………………………………..

31. How much do you like:

|  | **I like it a lot** | **I like it a bit** | **I don’t like it/**  **I don’t do it** |
| --- | --- | --- | --- |
| **Eating Asian food,**  **such as curry** |  |  |  |
| **Eating English food, such as fish and chips** |  |  |  |
| **Wearing Asian clothes such as**  **Salwar kameez or kurta** |  |  |  |
| **Wearing clothes**  **such as jeans,**  **t-shirts or dresses** |  |  |  |

32.

|  | **Yes** | **No** |
| --- | --- | --- |
| **Do you and your**  **family celebrate**  **Eid?** |  |  |
| **Do you and your**  **family celebrate**  **Christmas?** |  |  |

**Thank you for your time!**

Researcher/Teacher only:

Tick if the child was assisted to complete the questionnaire.

**Table S1: Vulnerability across domains of wellbeing**

**Table S1a: Response to items within ‘Home, Family and Family Relationships’ domain**

| **How often does your family get along well** | **Number** | **Percentage (95% CI)** |
| --- | --- | --- |
| Never | 729 | 4.8% (4.4%-5.1%) |
| Some of the time | 6,845 | 44.8% (44.1%-45.6%) |
| All of the time | 7,689 | 50.4% (49.6%-51.2%) |
| Missing | 378 |  |
| Total | 15,641 | 100.0% |
| **Does your home have a garden where you can play?** | **Number** | **Percentage (95% CI)** |
| No | 2,036 | 13.2% (12.7%-13.8%) |
| Yes | 13,357 | 86.8% (86.2%-87.3%) |
| Missing | 248 |  |
| Total | 15,641 | 100.0% |
| **Do you have a park near your home where you can play with your friends?** | **Number** | **Percentage (95% CI)** |
| No | 4,711 | 30.8% (30.1%-31.5%) |
| Yes | 10,584 | 69.2% (68.5%-69.9%) |
| Missing | 346 |  |
| Total | 15,641 | 100.0% |
| **How often do you play in a park?** | **Number** | **Percentage (95% CI)** |
| Never | 1,137 | 7.5% (7.1%-7.9%) |
| Sometimes | 11,701 | 76.9% (76.2%-77.5%) |
| Very often | 2,386 | 15.7% (15.1%-16.3%) |
| Missing | 417 |  |
| Total | 15,641 | 100.0% |

**Supplementary Table S1b: Response to items within ‘Material Resources’ domain**

| **Do you have a warm winter coat?** | **Number** | **Percentage (95% CI)** |
| --- | --- | --- |
| No | 1,402 | 10% (9.5%-10.5%) |
| Yes | 12,594 | 90% (89.5%-90.5%) |
| Missing | 1,645 |  |
| Total | 15,641 | 100.0% |
| **Do you have three meals every day?** | **Number** | **Percentage (95% CI)** |
| No | 2,243 | 15.9% (15.3%-16.5%) |
| Yes | 11,891 | 84.1% (83.5%-84.7%) |
| Missing | 1,507 |  |
| Total | 15,641 | 100.0% |
| **Do you have a computer/laptop/tablet with internet at home?** | **Number** | **Percentage (95% CI)** |
| No | 2,733 | 19.1% (18.5%-19.8%) |
| Yes | 11,563 | 80.9% (80.2%-81.5%) |
| Missing | 1,345 |  |
| Total | 15,641 | 100.0% |
| **How often do you worry about how much money your family has?** | **Number** | **Percentage (95% CI)** |
| Never | 5,264 | 34.1% (33.3%-34.8%) |
| Some of the time | 6,124 | 39.7% (38.9%-40.4%) |
| All of the time | 4,054 | 26.3% (25.6%-27%) |
| Missing | 199 |  |
| Total | 15,641 | 100.0% |

**Supplementary Table S1c: Response to items within ‘Friends and School’ domain**

| **How much do you like your school?** | **Number** | **Percentage (95% CI)** |
| --- | --- | --- |
| Don’t like it | 2,032 | 13.1% (12.6%-13.6%) |
| Like it a bit | 4,691 | 30.2% (29.5%-31%) |
| Like it a lot | 8,797 | 56.7% (55.9%-57.5%) |
| Missing | 121 |  |
| Total | 15,641 | 100.0% |
| **How many friends do you have?** | **Number** | **Percentage (95% CI)** |
| Not many | 2,257 | 14.5% (14%-15.1%) |
| Some | 3,647 | 23.5% (22.8%-24.2%) |
| Lots | 9,623 | 62% (61.2%-62.7%) |
| Missing | 114 |  |
| Total | 15,641 | 100.0% |
| **How often do other children bully you?** | **Number** | **Percentage (95% CI)** |
| Never | 7,276 | 47.3% (46.6%-48.1%) |
| Some of the time | 6,413 | 41.7% (41%-42.5%) |
| All of the time | 1,679 | 10.9% (10.4%-11.4%) |
| Missing | 273 |  |
| Total | 15,641 | 100.0% |
| **How often are you mean to other children at school?** | **Number** | **Percentage (95% CI)** |
| Never | 10,806 | 70.9% (70.2%-71.6%) |
| Some of the time | 3,645 | 23.9% (23.2%-24.6%) |
| All of the time | 794 | 5.2% (4.9%-5.6%) |
| Missing | 396 |  |
| Total | 15,641 | 100.0% |
| **How often do you feel left out by other children?** | **Number** | **Percentage (95% CI)** |
| Never | 5,598 | 36.2% (35.5%-37%) |
| Some of the time | 8,129 | 52.6% (51.8%-53.4%) |
| All of the time | 1,729 | 11.2% (10.7%-11.7%) |
| Missing | 185 |  |
| Total | 15,641 | 100.0% |

**Supplementary Table S1d: Response to items within ‘Subjective Wellbeing’ domain**

| **How often do you feel happy?** | **Number** | **Percentage (95% CI)** |
| --- | --- | --- |
| Never | 585 | 3.8% (3.5%-4.1%) |
| Some of the time | 8,306 | 54.2% (53.5%-55%) |
| All of the time | 6,420 | 41.9% (41.2%-42.7%) |
| Missing | 330 |  |
| Total | 15,641 | 100.0% |
| **How often do you feel sad?** | **Number** | **Percentage (95% CI)** |
| Never | 3,837 | 24.9% (24.2%-25.6%) |
| Some of the time | 10,715 | 69.5% (68.8%-70.2%) |
| All of the time | 863 | 5.6% (5.2%-6%) |
| Missing | 226 |  |
| Total | 15,641 | 100.0% |
| **How often are you ill or unwell?** | **Number** | **Percentage (95% CI)** |
| Never | 2,216 | 14.4% (13.8%-14.9%) |
| Some of the time | 11,966 | 77.6% (76.9%-78.2%) |
| All of the time | 1,247 | 8.1% (7.7%-8.5%) |
| Missing | 212 |  |
| Total | 15,641 | 100.0% |
| **Keep worries to self** | **Number** | **Percentage (95% CI)** |
| No | 10,655 | 68.8% (68.1%-69.5%) |
| Yes | 4,828 | 31.2% (30.5%-31.9%) |
| Missing | 158 |  |
| Total | 15,641 | 100.0% |
| **When I find something really hard I can work out what to do next** | **Number** | **Percentage (95% CI)** |
| Never | 1,299 | 8.6% (8.1%-9%) |
| Some of the time | 9,429 | 62.3% (61.5%-63.1%) |
| All of the time | 4,409 | 29.1% (28.4%-29.9%) |
| Missing | 504 |  |
| Total | 15,641 | 100.0% |

**Table S2: Percentage primary school children with zero, one, or more than one vulnerability in each domain of wellbeing**

| **Home, family, relationships** | **Number** | **Percentage (95% CI)** |
| --- | --- | --- |
| None | 8,085 | 55.9% (55%-56.7%) |
| One | 4,902 | 33.9% (33.1%-34.6%) |
| More than one | 1,489 | 10.3% (9.8%-10.8%) |
| Missing | 1,165 |  |
| Total | 15,641 | 100.0% |
| **Material resources** | **Number** | **Percentage (95% CI)** |
| None | 6,331 | 48.3% (47.5%-49.2%) |
| One | 4,763 | 36.4% (35.5%-37.2%) |
| More than one | 2,008 | 15.3% (14.7%-16%) |
| Missing | 2,539 |  |
| Total | 15,641 | 100.0% |
| **Friends and school** | **Number** | **Percentage (95% CI)** |
| None | 5,204 | 35% (34.3%-35.8%) |
| One | 6,189 | 41.7% (40.9%-42.5%) |
| More than one | 3,455 | 23.3% (22.6%-24%) |
| Missing | 793 |  |
| Total | 15,641 | 100.0% |
| **Subjective wellbeing** | **Number** | **Percentage (95% CI)** |
| None | 8,312 | 57.4% (56.6%-58.2%) |
| One | 4,676 | 32.3% (31.5%-33.1%) |
| More than one | 1,496 | 10.3% (9.8%-10.8%) |
| Missing | 1,157 |  |
| Total | 15,641 | 100.0% |

**Supplementary Table S3: Associations of gender with vulnerabilities in child wellbeing domains**

**Supplementary Table S3a – Home, family, family relations**

|  | **Family never get along well together**  (N = 15,263  non-missing) | | **No garden where they can play**  (N = 15,393  non-missing) | | **No park near home where they can play with friends**  (N = 15,295  non-missing) | | **Never plays**  **in park**  (N = 15,224  non-missing) | |
| --- | --- | --- | --- | --- | --- | --- | --- | --- |
| **Gender** | **N** | **Percentage (95% CI)** | **N** | **Percentage (95% CI)** | **N** | **Percentage (95% CI)** | **N** | **Percentage (95% CI)** |
| Female (N = 7,647) | 296 | 4.0%  (3.5%-4.4%) | 891 | 11.8%  (11.1%-12.5%) | 2225 | 11.8%  (11.1%-12.5%) | 456 | 6.1%  (5.6%-6.7%) |
| Male (N = 7,994) | 433 | 5.6%  (5.1%-6.1%) | 1145 | 14.6%  (13.8%-15.4%) | 2486 | 14.6%  (13.8%-15.4%) | 681 | 8.8%  (8.2%-9.4%) |

**Supplementary Table S3b – Material resources**

|  | **No warm**  **winter coat**  (N = 13,996  non-missing) | | **Not having three**  **meals a day**  (N = 14,134  non-missing) | | **No internet**  **at home**  (N = 14,296  non-missing) | | **Worried about**  **money all the time**  (N = 15,442  non-missing) | |
| --- | --- | --- | --- | --- | --- | --- | --- | --- |
| **Gender** | **N** | **Percentage (95% CI)** | **N** | **Percentage (95% CI)** | **N** | **Percentage (95% CI)** | **N** | **Percentage (95% CI)** |
| Female (N = 7,647) | 616 | 8.9%  (8.2%-9.6%) | 983 | 14.0%  (13.3%-14.9%) | 1383 | 19.7%  (18.8%-20.6%) | 1771 | 23.4%  (22.5%-24.4%) |
| Male (N = 7,994) | 786 | 11.1%  (10.4%-11.9%) | 1260 | 17.7%  (16.8%-18.6%) | 1350 | 18.6%  (17.7%-19.5%) | 2283 | 28.9%  (28.0%-30.0%) |

**Supplementary Table S3c – Friends and school**

|  | **Does not like**  **school**  (N = 15,520  non-missing) | | **Not many**  **friends**  (N = 15,527  non-missing) | | **Bullied some or all of the time**  (N = 15,368  non-missing) | | **Mean to others all the time**  (N = 15,245  non-missing) | | **Feel left out all**  **the time**  (N = 15,456  non-missing) | |
| --- | --- | --- | --- | --- | --- | --- | --- | --- | --- | --- |
| **Gender** | **N** | **Percentage (95% CI)** | **N** | **Percentage (95% CI)** | **N** | **Percentage (95% CI)** | **N** | **Percentage (95% CI)** | **N** | **Percentage (95% CI)** |
| Female (N = 7,647) | 606 | 8.0%  (7.4%-8.6%) | 1269 | 16.7%  (15.9%-17.6%) | 4192 | 55.8%  (54.7%-56.9%) | 252 | 3.4%  (3.0%-3.8%) | 916 | 12.1%  (11.4%-12.8%) |
| Male (N = 7,994) | 1426 | 18.0%  (17.1%-18.8%) | 988 | 12.5%  (11.8%-13.2%) | 3900 | 49.7%  (48.6%-50.8%) | 542 | 7.0%  (6.4%-7.6%) | 813 | 10.3%  (9.7%-11.0%) |

**Supplementary Table S3d – Subjective wellbeing**

|  | **Never happy**  (N = 15,331  non-missing) | | **Always sad**  (N = 15,415  non-missing) | | **Always ill or unwell**  (N = 15,429  non-missing) | | **Keeps worries**  **to self**  (N = 15,483  non-missing) | | **Cannot work out what to do when things are hard**  (N = 15,137  non-missing) | |
| --- | --- | --- | --- | --- | --- | --- | --- | --- | --- | --- |
| **Gender** | **N** | **Percentage (95% CI)** | **N** | **Percentage (95% CI)** | **N** | **Percentage (95% CI)** | **N** | **Percentage (95% CI)** | **N** | **Percentage (95% CI)** |
| Female (N = 7,647) | 179 | 2.4%  (2.1%-2.8%) | 412 | 5.5%  (5.0%-6.0%) | 581 | 7.7%  (7.1%-8.3%) | 2166 | 28.5%  (27.5%-29.6%) | 555 | 7.5%  (6.9%-8.1%) |
| Male (N = 7,994) | 406 | 5.2%  (4.7%-5.7%) | 451 | 5.7%  (5.2%-6.3%) | 666 | 8.5%  (7.9%-9.1%) | 2662 | 33.7%  (32.7%-34.8%) | 744 | 9.6%  (9.0%-10.3%) |

**Supplementary Table S3e: Associations of gender for each child wellbeing domain**

|  | **Home, family and family relationships**  (N = 14,476 non-missing) | | **Material resources**  (N = 13,102 non-missing) | | **Friends and school**  (N = 14,848 non-missing) | | **Subjective wellbeing**  (N = 14,484 non-missing) | |
| --- | --- | --- | --- | --- | --- | --- | --- | --- |
| **Gender** | **N** | **Percentage**  **(95% CI)** | **N** | **Percentage**  **(95% CI)** | **N** | **Percentage**  **(95% CI)** | **N** | **Percentage**  **(95% CI)** |
| Female (N = 7,647) | 7,117 | 41.7%  (40.6%-42.9%) | 6,520 | 48.8%  (47.6%-50.0%) | 7,284 | 65.0%  (63.9%-66.1%) | 7,148 | 39.5%  (38.4%-40.7%) |
| Male (N = 7,994) | 7,359 | 46.5%  (45.3%-47.6%) | 6,582 | 54.5%  (53.3%-55.7%) | 7,564 | 64.9%  (63.8%-66.0%) | 7,336 | 45.6%  (44.5%-46.7%) |

**Supplementary Table S4: Associations of ethnicity with vulnerabilities in child wellbeing domains**

**Supplementary Table S4a – Home, family, family relations**

|  | **Family never get along well together**  (N = 15,263  non-missing) | | **No garden where they can play**  (N = 15,393  non-missing) | | **No park near home where they can play with friends**  (N = 15,295  non-missing) | | **Never plays**  **in park**  (N = 15,224  non-missing) | |
| --- | --- | --- | --- | --- | --- | --- | --- | --- |
| **Ethnicity** | **N** | **Percentage (95% CI)** | **N** | **Percentage (95% CI)** | **N** | **Percentage**  **(95% CI)** | **N** | **Percentage (95% CI)** |
| Pakistani (N = 7,031) | 278 | 4.1%  (3.6%-4.6%) | 837 | 12.1%  (11.4%-12.9%) | 2243 | 32.7%  (31.6%-33.9%) | 470 | 6.9%  (6.3%-7.5%) |
| White British (N = 4,247) | 265 | 6.3%  (5.6%-7.1%) | 562 | 13.4%  (12.4%-14.5%) | 1247 | 29.9%  (28.6%-31.3%) | 374 | 9.0%  (8.2%-9.9%) |
| Mixed (N = 900) | 55 | 6.3%  (4.9%-8.1%) | 134 | 15.0%  (12.8%-17.5%) | 254 | 28.7%  (25.8%-31.8%) | 75 | 8.5%  (6.8%-10.5%) |
| White Other (N = 707) | 24 | 3.5%  (2.3%-5.2%) | 125 | 18.0%  (15.3%-21%) | 188 | 27.1%  (23.9%-30.6%) | 46 | 6.7%  (5.1%-8.9%) |
| Bangladeshi (N = 471) | 18 | 3.9%  (2.5%-6.2%) | 73 | 15.8%  (12.8%-19.5%) | 154 | 33.6%  (29.4%-38.0%) | 32 | 6.9%  (4.9%-9.6%) |
| Indian (N = 357) | 17 | 4.9%  (3.0%-7.7%) | 64 | 18.1%  (14.4%-22.4%) | 121 | 34.4%  (29.6%-39.5%) | 25 | 7.2%  (4.9%-10.4%) |
| Black/ Black British (N = 277) | 5 | 1.8%  (0.8%-4.3%) | 49 | 18.0%  (13.9%-23%) | 62 | 22.7%  (18.1%-28.1%) | 19 | 7.0%  (4.5%-10.7%) |
| Gypsy/ Irish traveller (N = 190) | 13 | 7.3%  (4.3%-12.2%) | 28 | 15.2%  (10.7%-21.2%) | 54 | 29.2%  (23.1%-36.1%) | 18 | 9.9%  (6.3%-15.2%) |
| Other (N = 425) | 14 | 3.4%  (2.0%-5.7%) | 76 | 18.1%  (14.7%-22.1%) | 129 | 31.5%  (27.2%-36.1%) | 35 | 8.5%  (6.1%-11.5%) |

**Supplementary Table S4b – Material resources**

|  | **No warm**  **winter coat**  (N = 13,996  non-missing) | | **Not having three**  **meals a day**  (N = 14,134  non-missing) | | **No internet**  **at home**  (N = 14,296  non-missing) | | **Worried about**  **money all the time**  (N = 15,442  non-missing) | |
| --- | --- | --- | --- | --- | --- | --- | --- | --- |
| **Ethnicity** | **N** | **Percentage (95% CI)** | **N** | **Percentage (95% CI)** | **N** | **Percentage (95% CI)** | **N** | **Percentage**  **(95% CI)** |
| Pakistani (N = 7,031) | 609 | 9.8%  (9.1%-10.6%) | 1116 | 17.9%  (16.9%-18.8%) | 1141 | 18.0%  (17.0%-18.9%) | 2031 | 29.3%  (28.2%-30.3%) |
| White British (N = 4,247) | 432 | 11.1%  (10.1%-12.1%) | 470 | 11.9%  (10.9%-12.9%) | 880 | 22.3%  (21.0%-23.6%) | 912 | 21.6%  (20.4%-22.9%) |
| Mixed (N = 900) | 76 | 9.6%  (7.8%-11.9%) | 145 | 18.0%  (15.5%-20.8%) | 159 | 19.7%  (17.1%-22.6%) | 223 | 25.3%  (22.5%-28.3%) |
| White Other (N = 707) | 55 | 8.9%  (6.9%-11.4%) | 90 | 14.4%  (11.8%-17.3%) | 113 | 17.7%  (14.9%-20.8%) | 204 | 29.4%  (26.1%-32.9%) |
| Bangladeshi (N = 471) | 31 | 7.5%  (5.3%-10.5%) | 65 | 16.1%  (12.8%-20.0%) | 74 | 17.6%  (14.2%-21.5%) | 128 | 27.6%  (23.7%-31.8%) |
| Indian (N = 357) | 31 | 9.6%  (6.8%-13.3%) | 67 | 20.4%  (16.4%-25.1%) | 47 | 14.3%  (10.9%-18.5%) | 88 | 24.9%  (20.7%-29.7%) |
| Black/ Black British (N = 277) | 20 | 8.0%  (5.2%-12.1%) | 40 | 15.9%  (11.9%-20.9%) | 36 | 13.7%  (10%-18.4%) | 74 | 26.8%  (21.9%-32.3%) |
| Gypsy/ Irish traveller (N = 190) | 15 | 9.4%  (5.7%-15%) | 32 | 19.8%  (14.3%-26.6%) | 43 | 25.3%  (19.3%-32.4%) | 73 | 39.0%  (32.3%-46.2%) |
| Other (N = 425) | 33 | 8.9%  (6.4%-12.2%) | 56 | 14.9%  (11.6%-18.8%) | 81 | 21.3%  (17.5%-25.7%) | 101 | 24.4%  (20.5%-28.8%) |

**Supplementary Table S4c – Friends and school**

|  | **Does not like**  **school**  (N = 15,520  non-missing) | | **Not many**  **friends**  (N = 15,527  non-missing) | | **Bullied some or all of the time**  (N = 15,368  non-missing) | | **Mean to others all the time**  (N = 15,245  non-missing) | | **Feel left out all**  **the time**  (N = 15,456  non-missing) | |
| --- | --- | --- | --- | --- | --- | --- | --- | --- | --- | --- |
| **Ethnicity** | **N** | **Percentage (95% CI)** | **N** | **Percentage (95% CI)** | **N** | **Percentage (95% CI)** | **N** | **Percentage (95% CI)** | **N** | **Percentage (95% CI)** |
| Pakistani (N = 7,031) | 889 | 12.7%  (12.0%-13.5%) | 1029 | 14.7%  (13.9%-15.6%) | 3630 | 52.6%  (51.4%-53.7%) | 400 | 5.8%  (5.3%-6.4%) | 790 | 11.4%  (10.6%-12.1%) |
| White British (N = 4,247) | 652 | 15.4%  (14.4%-16.6%) | 662 | 15.7%  (14.6%-16.8%) | 2253 | 53.8%  (52.3%-55.3%) | 167 | 4.0%  (3.5%-4.7%) | 486 | 11.6%  (10.6%-12.6%) |
| Mixed (N = 900) | 128 | 14.4%  (12.2%-16.8%) | 113 | 12.7%  (10.6%-15%) | 459 | 51.9%  (48.6%-55.1%) | 41 | 4.7%  (3.5%-6.3%) | 98 | 11.0%  (9.1%-13.2%) |
| White Other (N = 707) | 69 | 9.8%  (7.8%-12.3%) | 106 | 15.2%  (12.7%-18%) | 328 | 47.1%  (43.4%-50.8%) | 44 | 6.4%  (4.8%-8.5%) | 74 | 10.6%  (8.5%-13.1%) |
| Bangladeshi (N = 471) | 54 | 11.5%  (8.9%-14.8%) | 53 | 11.3%  (8.7%-14.5%) | 235 | 51.1%  (46.5%-55.6%) | 24 | 5.2%  (3.5%-7.6%) | 40 | 8.5%  (6.3%-11.4%) |
| Indian (N = 357) | 33 | 9.3%  (6.7%-12.8%) | 46 | 13.0%  (9.9%-16.9%) | 183 | 52.0%  (46.8%-57.2%) | 15 | 4.3%  (2.6%-7.0%) | 31 | 8.8%  (6.3%-12.3%) |
| Black/ Black British (N = 277) | 32 | 11.6%  (8.3%-15.9%) | 23 | 8.3%  (5.6%-12.2%) | 144 | 52.9%  (47%-58.8%) | 14 | 5.2%  (3.1%-8.5%) | 24 | 8.8%  (6.0%-12.8%) |
| Gypsy/ Irish traveller (N = 190) | 20 | 10.7%  (7.0%-16.0%) | 30 | 16.0%  (11.4%-21.9%) | 79 | 42.2%  (35.4%-49.4%) | 18 | 9.9%  (6.3%-15.2%) | 21 | 11.3%  (7.5%-16.7%) |
| Other (N = 425) | 53 | 12.6%  (9.8%-16.2%) | 60 | 14.3%  (11.3%-18%) | 233 | 56.3%  (51.5%-61.0%) | 24 | 5.8%  (3.9%-8.5%) | 40 | 9.6%  (7.1%-12.8%) |

**Supplementary Table S4d – Subjective wellbeing**

|  | **Never happy**  (N = 15,331  non-missing) | | **Always sad**  (N = 15,415  non-missing) | | **Always ill or unwell**  (N = 15,429  non-missing) | | **Keeps worries**  **to self**  (N = 15,483  non-missing) | | **Cannot work out what to do when things are hard**  (N = 15,137  non-missing) | |
| --- | --- | --- | --- | --- | --- | --- | --- | --- | --- | --- |
| **Ethnicity** | **N** | **Percentage (95% CI)** | **N** | **Percentage (95% CI)** | **N** | **Percentage (95% CI)** | **N** | **Percentage (95% CI)** | **N** | **Percentage (95% CI)** |
| Pakistani (N = 7,031) | 281 | 4.1%  (3.7%-4.6%) | 406 | 5.9% (  5.3%-6.4%) | 622 | 9%  (8.3%-9.7%) | 2222 | 31.9%  (30.8%-33%) | 570 | 8.4%  (7.7%-9.1%) |
| White British (N = 4,247) | 168 | 4%  (3.5%-4.7%) | 223 | 5.3%  (4.7%-6%) | 305 | 7.2%  (6.5%-8.1%) | 1268 | 30.2%  (28.8%-31.6%) | 372 | 9%  (8.2%-9.9%) |
| Mixed (N = 900) | 38 | 4.3%  (3.2%-5.9%) | 55 | 6.2%  (4.8%-8%) | 73 | 8.3%  (6.6%-10.3%) | 281 | 31.5%  (28.6%-34.7%) | 88 | 10.1%  (8.2%-12.2%) |
| White Other (N = 707) | 26 | 3.8%  (2.6%-5.5%) | 43 | 6.2%  (4.6%-8.2%) | 50 | 7.2%  (5.5%-9.4%) | 210 | 30%  (26.8%-33.5%) | 69 | 10.1%  (8.1%-12.7%) |
| Bangladeshi (N = 471) | 14 | 3.1%  (1.8%-5.1%) | 21 | 4.5%  (3%-6.9%) | 45 | 9.7%  (7.3%-12.8%) | 139 | 29.8%  (25.8%-34.1%) | 22 | 4.8%  (3.2%-7.1%) |
| Indian (N = 357) | 6 | 1.7%  (0.8%-3.8%) | 20 | 5.7%  (3.7%-8.7%) | 22 | 6.3%  (4.2%-9.4%) | 112 | 31.6%  (27%-36.7%) | 23 | 6.7%  (4.5%-9.9%) |
| Black/ Black British (N = 277) | 6 | 2.2%  (1%-4.8%) | 16 | 5.8%  (3.6%-9.3%) | 19 | 6.9%  (4.4%-10.5%) | 79 | 28.6%  (23.6%-34.2%) | 15 | 5.6%  (3.4%-9.1%) |
| Gypsy/ Irish traveller (N = 190) | 6 | 3.3%  (1.5%-7.1%) | 13 | 7%  (4.1%-11.7%) | 16 | 8.6%  (5.3%-13.5%) | 60 | 32.3%  (25.9%-39.3%) | 27 | 15.2%  (10.6%-21.2%) |
| Other (N = 425) | 14 | 3.4%  (2%-5.6%) | 23 | 5.5%  (3.7%-8.2%) | 36 | 8.6%  (6.2%-11.7%) | 135 | 32.5%  (28.1%-37.1%) | 38 | 9.3%  (6.8%-12.5%) |

**Supplementary Table S7: Associations of Index of Multiple Deprivation for each child wellbeing domain**

| **IMD 2019 Quintile (1 = most materially deprived)** | **Home, family and family relationships**  (N = 5,703 non-missing) | | **Material resources**  (N = 5,153 non-missing) | | **Friends and school**  (N = 5,869 non-missing) | | **Subjective wellbeing**  (N = 5,706 non-missing) | |
| --- | --- | --- | --- | --- | --- | --- | --- | --- |
|  | **N** | **Percentage (95% CI)** | **N** | **Percentage (95% CI)** | **N** | **Percentage (95% CI)** | **N** | **Percentage (95% CI)** |
| 1 (N = 4,334) | 1817 | 45.4%  (43.8%-46.9%) | 1907 | 53.5%  (51.9%-55.1%) | 2713 | 65.7%  (64.2%-67.1%) | 1726 | 43.1%  (41.6%-44.6%) |
| 2 (N = 1,303) | 511 | 42.0%  (39.2%-44.8%) | 546 | 48.5%  (45.6%-51.5%) | 797 | 64.0%  (61.3%-66.6%) | 482 | 39.5%  (36.8%-42.3%) |
| 3 (N = 308) | 103 | 35.5%  (30.0%-41.0%) | 119 | 43.9%  (38.0%-49.8%) | 182 | 61.1%  (55.5%-66.6%) | 113 | 39.6%  (34.0%-45.3%) |
| 4 (N = 139) | 50 | 38.5%  (30.1%-46.8%) | 49 | 37.1%  (28.9%-45.4%) | 80 | 60.6%  (52.3%-68.9%) | 47 | 35.1%  (27.0%-43.2%) |
| 5 (N = 59) | 23 | 39.0%  (26.5%-51.4%) | 19 | 33.9%  (21.5%-46.3%) | 33 | 56.9%  (44.2%-69.6%) | 19 | 32.8%  (20.7%-44.8%) |
